# A disease concept model for *STXBP1*-related disorders

**DOI:** 10.1101/2022.08.05.22278197

**Authors:** Katie R Sullivan, Sarah M Ruggiero, Julie Xian, Kim M Thalwitzer, Sydni Stewart, Mahgenn Cosico, Jackie Steinberg, James Goss, Anna Pfalzer, Kyle J Horning, Nicole Weitzel, Sydney Corey, Laura Conway, Charlene Son Rigby, Terry Jo Bichell, Ingo Helbig

**Author notes:** Corresponding author: **Ingo Helbig, MD**, Corresponding author’s address: Division of Neurology, Children’s Hospital of Philadelphia, Philadelphia, PA 19104, Corresponding author’s, Corresponding author’s. Jackie Steinburg j.

## Abstract

**Objective:** *STXBP1*-related disorders are common genetic epilepsies and neurodevelopmental disorders, but the impact of symptoms across clinical domains is poorly understood. Disease concept models are formal frameworks to assess the lived experience of individuals and their families and provide a basis for generating outcome measures.

**Methods:** We conducted semi-structured, qualitative interviews with 19 caregivers of 16 individuals with *STXBP1*-related disorders and 7 healthcare professionals. We systematically coded themes using NVivo software and grouped concepts into the domains of symptoms, symptom impact and caregiver impact. We quantified the frequency of concepts throughout the life span and across clinical subgroups stratified by seizure history and developmental trajectories.

**Results:** Over 25 hours of interviews, we coded a total of 3626 references to 38 distinct concepts. In addition to well recognized clinical features such as developmental delay (n=240 references), behavior (n=201), and seizures (n=147), we identified previously underrepresented symptoms including gastrointestinal (n=68) and respiratory symptoms (n=24) and pain (n=30). The most frequently referenced symptom impacts were autonomy (n=96), socialization (n=64), and schooling (n=61). Emotional impact (n=354), support (n=200), and daily life & activities (n=108) were highly cited caregiver impacts. We found that seizures (OR=8.84, 95% CI 5.97-13.1) were more commonly referenced in infancy than in other age groups, while behavior (OR=2.15, 95% CI 1.56-2.95), and socialization (OR=2.26, 95% CI 1.28-3.96) were more likely to be referred in childhood. We found that caregivers of individuals with ongoing seizures were less likely to reference developmental delay (OR=0.59, 95% CI 0.42-0.82), possibly due to the relatively high impact of seizures (OR=8.84, 95% CI 5.97-13.1).

**Significance:** *STXBP1*-related disorders are complex conditions affecting a wide range of clinical and social domains. We comprehensively mapped symptoms and their impact on families to generate a comprehensive disease model as a foundation for clinical endpoints in future trials.

**Key points:**

- Disease concept models (DCMs) are formal frameworks that capture the relationship between symptoms, concerns, and impact on daily life based on qualitative interviews
- Endpoints for future clinic studies involving *STXBP1*-related disorder need to be relevant to patients and families
- Interviewing a diverse cohort of caregivers and healthcare professionals allows DCMs to be generalizable and reveals high priority & novel disease concepts for treatment
- Disease concepts can vary depending on an affected individual’s age, necessitating longitudinal record of concepts
- Disease concepts can differ across clinical subgroups stratified by epilepsy and developmental histories making inclusion of variable disorder presentations important

## Introduction

*STXBP1*-related disorders are common developmental and epileptic encephalopathies (DEEs), a group of early-onset epilepsies and neurodevelopmental disorders [1-3]. First described in 2008 in a cohort of individuals with Ohtahara syndrome [4], *STXBP1*-related disorders are now known to present with a broader range of clinical features and neurodevelopmental trajectories [3, 5-7]. While largely unknown only several years ago, remarkable progress has been made towards understanding the clinical spectrum and longitudinal disease trajectory in individuals with *STXBP1*-related disorders. We previously identified emerging genotype-phenotype relationships within a larger cohort of 534 individuals, identifying several clinical features associated with specific variant types. For example, protein-truncating variants including stop codon mutations, splice-site variants, and frameshift variants were associated with a three-fold increased risk of infantile spasms, while the recurrent p.R406C/H variant was associated with a two-fold increased risk for spasticity and suppression-burst EEG [3]. In addition, the adult phenotype [8] and the interaction between early seizures and development are increasingly being investigated [9]. Furthermore, significant progress has been made towards disease-specific therapies within the last two years, including chemical chaperones [10] and novel avenues for targeted gene therapies and precision medicine approaches.

However, little is known about the full impact of *STXBP1*-related disorders on affected individuals and their caregivers, an understand of which is critical identify outcomes of importance to patients and inform optimal windows for interventions. Disease concept models (DCMs) are formal frameworks that capture the lived experience of a disorder based on qualitative interviews. While clinical studies are often limited to pre-defined symptoms such as seizures or achievement of developmental milestones, DCMs use an open-ended approach to capture the entirety of the features associated with a given condition. Within frameworks for drug development, regulators such as the US Food and Drug Administration (FDA) and the European Medicines Agency (EMA) increasingly rely on formal DCMs to prioritize outcomes for clinical trials [11-13]. DCMs have already been developed for Angelman syndrome and Dravet syndrome, leading to significant progression in understanding the overall impact of clinical features beyond epilepsy and neurodevelopmental symptoms in these disorders [14, 15].

Here, we develop a DCM for *STXBP1*-related disorders based on formal qualitative interviews with caregivers and healthcare professionals revealing 38 concepts, spanning symptoms, symptom impacts, and caregiver impact domains. We demonstrate a dynamic landscape of symptoms and their impact on families, capturing subgroups stratified by seizure history and developmental trajectories across the age-span, providing insight for future natural history studies and trial design.

## Methods

### Study inclusion of caregivers and healthcare providers

Caregivers of individuals with *STXBP1*-related disorders were recruited through the Epilepsy Neurogenetics Initiative (ENGIN) and the Epilepsy Genetics Research Project (EGRP) at Children’s Hospital of Philadelphia (CHOP) and the *STXBP1* Foundation. To ensure a diverse patient population, we included caregivers of individuals with *STXBP1*-related disorders of different ages, variable clinical spectrums regarding seizure histories, developmental trajectories, demographic features, and family living situations. Healthcare professionals were recruited through snowball sampling.

### Framework conceptualization and interview process

Semi-structured interviews were conducted virtually based on a formally developed framework consisting of open-ended questions (**Supplemental File 1-7**). An inductive thematic analysis approach was used to identify concepts described by participants rather than defined *a priori* [16]. Transcripts were analyzed using NVivo, a HIPAA-approved qualitative data analysis computer software. Coding was performed based on a formally developed code book consisting of specific concepts, which were categorized in the domains: symptoms, symptom impacts, and caregiver impacts (**Supplemental File 8**). For example, concepts, including references to behavior, seizures, and tone were classified as symptoms, while schooling, socialization, and daily life & activities were defined as symptom impacts. Two members of the study team coded in parallel and comparative methods were used to ensure consistency [17].

We assessed the frequency of references to certain concepts across all interviews. To elucidate age-related impacts of *STXBP1*-related disorders, we measured concepts within defined life spans including infancy (<12 months), toddlerhood (1-4 years), childhood (5-12 years), teenage & adolescence (13-17 years) and adulthood (18+ years). We also compared concepts across clinical subgroups of seizure history and ability of independent ambulation and verbal communication. One individual, who was under one year of age at the time of study inclusion, was excluded from these subgroup-analyses.

### Assessment of concept saturation and statistical analysis

Concept saturation is defined as the point in the collection of qualitative data when new data no longer adds to the body of knowledge. To properly reach saturation, sampling must be widely inclusive for all categories which might influence qualitative responses. Concept saturation was assessed using a data saturation matrix generated in NVivo Qualitative Software. Concept saturation was achieved with 16 interviews, suggesting an adequate sample size. When stratifying by *STXBP1*-related disorder symptoms, references were normalized to the total number of children within each subgroup. Statistical analysis was performed using R statistical framework to determine associations between concepts. Statistical significance was determined using Fisher’s exact test, and correction for multiple testing was performed using false discovery rate of 5%. Findings are presented as nominal associations with odds ratios and 95% confidence intervals.

## Results

### Generation of a disease concept model identified 38 concepts

We developed a semi-structured interview guide using a collaborating organizations’ preliminary *STXBP1*-related disorder disease concept model (DCM) and published interview guides from Dravet syndrome [14]. Disease features were reviewed and tailored to individuals with *STXBP1*-related disorders and their families based on current literature and the study team’s clinical experience in caring for 101 unique individuals with *STXBP1*-related disorders. The resulting interview guide consisted of 36 open questions, leading to conversations achieving both breadth and depth (**Supplemental File 1-7, Supplemental Table 1**). We transcribed a total of 25 hours and 23 minutes of interview time, resulting in 3626 references within 38 concepts and achieved an overall intercoder reliability of 88.2%.

### Interviewed caregivers and healthcare providers represented a wide demographic spectrum

The interview process included 19 caregivers and seven health care providers. Caregivers, including eight male participants, had a mean age of 42.5 years and cared for a total of 16 individuals with *STXBP1*-related disorders (**Table 1**). Additional demographic features included participants in same-sex couples, parental *STXBP1* pathogenic variant mosaicism, and households that speak English as a second language. We interviewed seven health care providers including two physicians, one genetic counselor, two physical therapists, one occupational therapist, and one educator.

**Table 1.** Participant demographics and clinical subgroups.

| Caregivers (n=19) |  | Healthcare providers (n=7) |  |
| --- | --- | --- | --- |
| Age (years) |  | Age (years) |  |
| Mean | 42.5 | Mean | 47.9 |
| Min, max | 33, 64 | Min, max | 28, 70 |
| Gender, n (%) |  | Gender, n (%) |  |
| Male | 8 (42%) | Male | 3 (42.9%) |
| Female | 11 (58%) | Female | 4 (57.1%) |
| Race, n (%) |  | Clinical specialty, n |  |
| White | 16 (84%) | Physician | 2 |
| Non-white | 3 (16%) | Physical therapist | 1 |
| Ethnicity, n (%) |  | Educator | 2 |
| Hispanic | 4 (21%) | Genetic Counselor | 1 |
| Non-Hispanic | 15 (79%) | Occupational Therapist | 1 |
| Individuals with *STXBP1*-related disorders (n=16) |  |  |  |
| Age at interview (years), n (%) |  | Race, n (%) |  |
| <1 | 1 (6%) | White | 13 (81%) |
| 1-5 | 6 (38%) | Non-white | 3 (19%) |
| 6-9 | 3 (19%) | Ethnicity, n (%) |  |
| 10-13 | 2 (13%) | Hispanic | 4 (25%) |
| 14-17 | 0 (0%) | Non-Hispanic | 12 (75%) |
| 18+ | 4 (25%) | Seizures (n=16), n (%) |  |
| Age of diagnosis (years), n (%) |  | No Seizures | 3 (19%) |
| <1 | 7 (44%) | Seizures | 13 (81%) |
| 1-5 | 4 (25%) | Past seizures | 8 (62%) |
| 6-9 | 2 (13%) | Current seizures | 5 (38%) |
| 10-13 | 0 (0%) | Development (n=15), n (%) |  |
| 14-17 | 1 (6%) | Global DD | 15 (100%) |
| 18+ | 2 (13%) | Ambulation (n=15), n (%) |  |
| Years since diagnosis |  | Non-ambulatory | 5 (33%) |
| Mean | 4.78 | Ambulatory | 10 (67%) |
| Min, max | 0.31, 11 | Verbal Communication (n=15), n (%) |  |
| Gender, n (%) |  | Verbal/some words | 3 (20%) |
| Male | 7 (44%) | Non-verbal | 12 (80%) |
| Female | 9 (60%) |  |  |

### Individuals with *STXBP1*-related disorders largely represent a pediatric patient population

We included seven male and nine female individuals with *STXBP1*-related disorders with a median age of 7.5 years with the youngest individual less than six months old and the oldest being more than 25 years old **(Table 1)**. This cohort includes four adult individuals but largely represents a pediatric patient population. The mean years since *STXBP1*-related disorders diagnosis was 4.8 with a minimum and maximum of 0.3 and 11 years respectively. Seven individuals in our cohort (43.75%) were diagnosed with *STXBP1*-related disorders within the first six months of life after the onset of seizures, which is consistent with the critical periods of seizure onset in *STXBP1*-related disorders and availability of clinical genetic testing. Outside of the first year of life, the average age of diagnosis was 8.8 years which included individuals with and without seizure histories. All but three individuals (n=13/16, 81.3%) had a history of seizures including five individuals with ongoing seizures at time of the interview. These individuals were considered in the ‘current seizures’ group and individuals with seizure remission in the ‘past seizures’ group. All individuals in this cohort experienced global developmental delay, 10 out of 15 children (66.6%) achieved independent movement, and three out of 15 children (20%) had some verbal expressive language or words.

### Symptoms (Domain 1): Most frequently reported symptoms by caregivers included developmental delay, behavior, and seizures

Developmental delay, behavior, and seizures were mentioned across all caregiver interviews **(Figure 1 & 5)**. We found developmental delay was referenced 240 times throughout the interviews and was frequently double coded with other concepts, such as: medical interaction (general medical; n=92), gross motor skills (n=58), expressive communication (n=56), and impacts to autonomy for the child with *STXBP1*-related disorders (n=52, **Figure 2)**. Caregivers referenced behavioral symptoms 201 times, categorizing them as negative in 61%, positive in 23%, and neutral in 17% of cases **(Supplementary Figure 1)**. Behavioral symptoms were frequently referenced with medical interaction (n=42), caregiver emotional impacts (n=41), caregiver impacts to daily life & activities (n=34), and developmental delays (n=34). Seizures were referenced 147 times and often mentioned in the context of medical interaction (n=120), emotional caregiver impacts (n=55), impact to the child’s sleep (n=15), and caregiver impacts to support (n=12).

**Figure 1.**
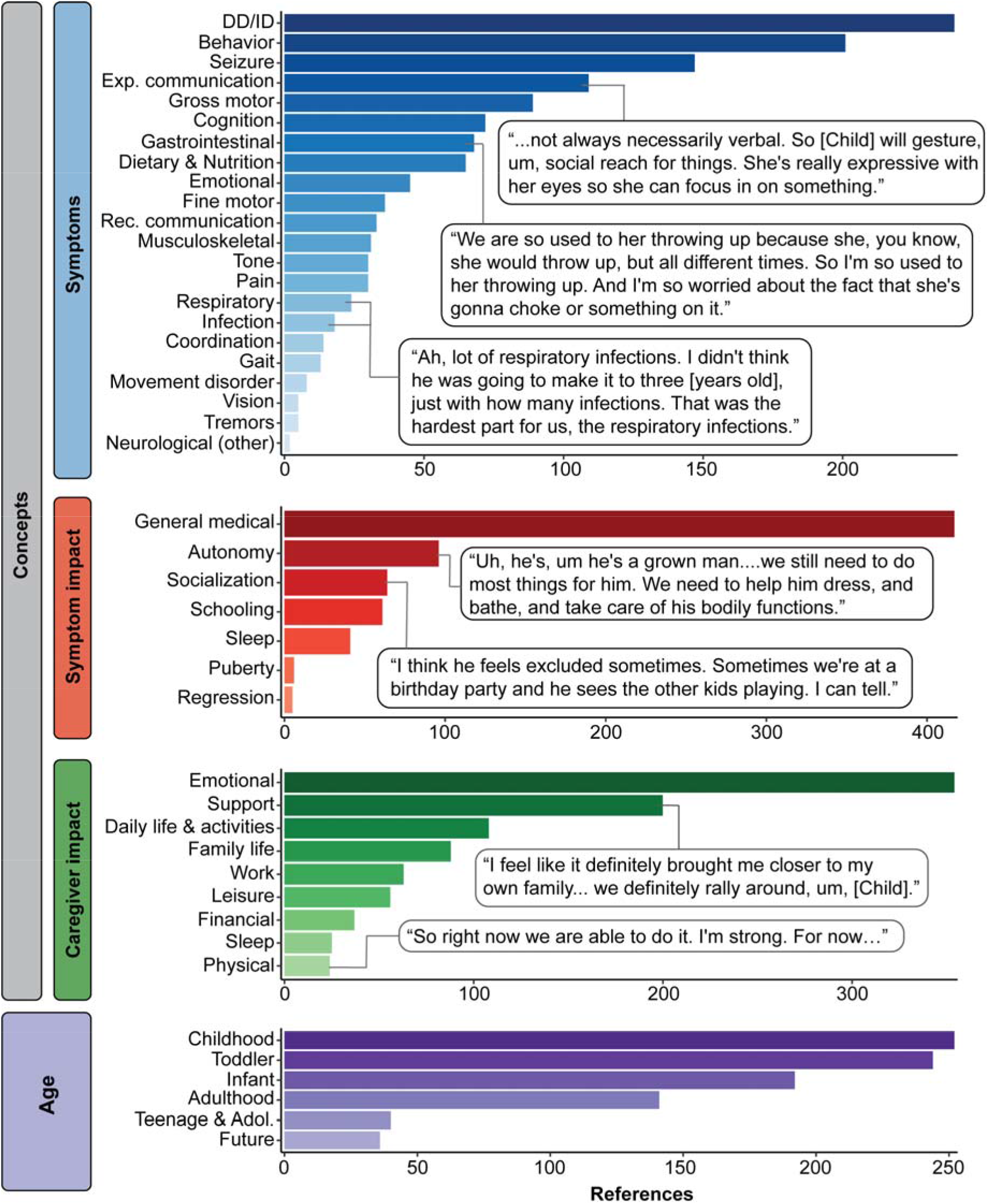
Disease concept model of *STXBP1*-related disorders. A total of 3,626 references across 19 caregivers were made referring to 38 distinct concepts categorized in the domains: symptoms, symptom impact, caregiver impacts, and across the across the life span (infancy: <12 months, toddlerhood: 1-4 years, childhood: 5-12 years, teenage & adolescence: 13-17 years, and adulthood 18+ years). In the domain for symptoms, expressive communication and receptive communication are notated as “exp. communication” and “rec. communication”, respectively.

**Figure 2.**
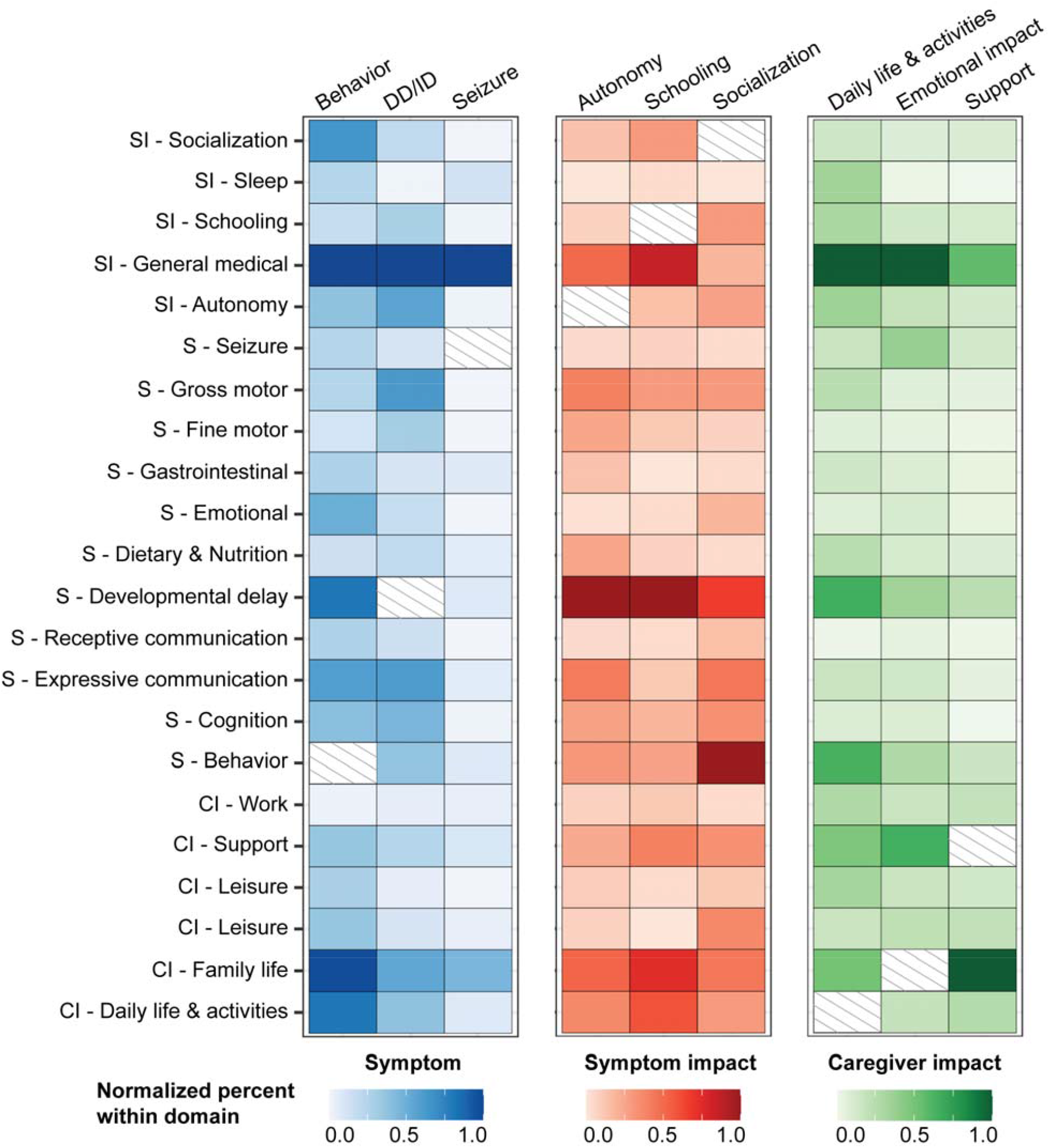
Distribution of co-codes among the most referenced concepts in each domain. Behavior, developmental delay or intellectual disability (DD/ID), and seizures were the most common symptoms, while autonomy, schooling, and socialization were the most common symptom impacts. The most frequent caregiver impacts were daily life & activities, emotional impact, and support. Most common co-coded concepts in each domain are shown, normalized in relation to the most frequently co-coded concept.

**Figure 3.**
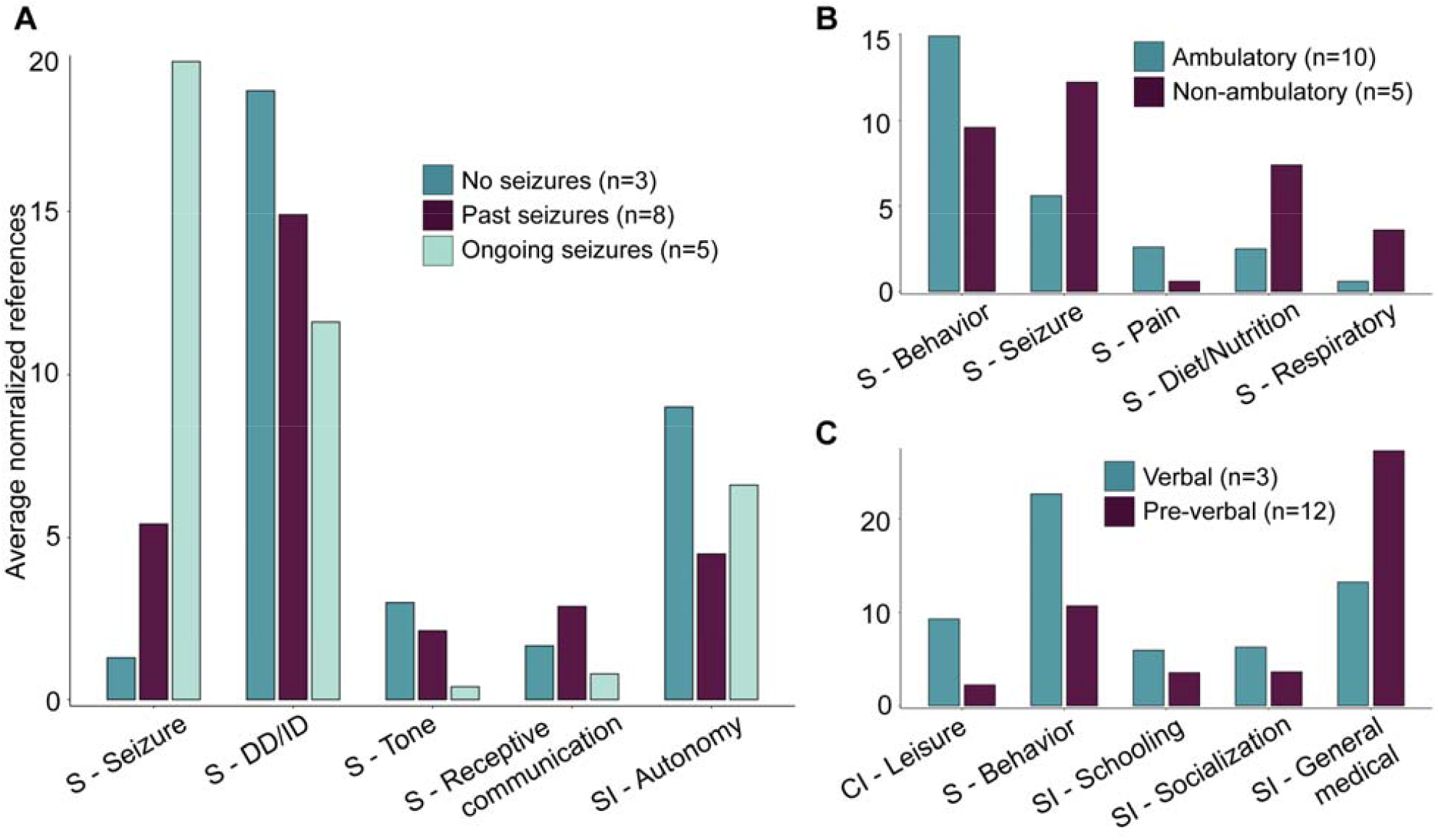
Analysis of subgroups stratified by seizure histories and developmental trajectories. Highlighted are the concepts that were significant across subgroups after correction for multiple testing with a false discovery rate of 5%. Height of each bar indicates the average references of concepts normalized to the number of individuals in each respective subgroup. **(A)** Differences in concepts across individuals without any history of seizures (n=3), individuals in seizure remission (n=8), and individuals with ongoing seizures (n=5). **(B)** Differences in concepts across individuals who did not achieve independent ambulation at the time of study inclusion (n=5) compared to who achieved independent ambulation (n=10). **(C)** Differences in concepts across individuals who did not use words to communicate (non-verbal, n=12) compared to those who used words to communicate (verbal, n=3).

**Figure 4.**
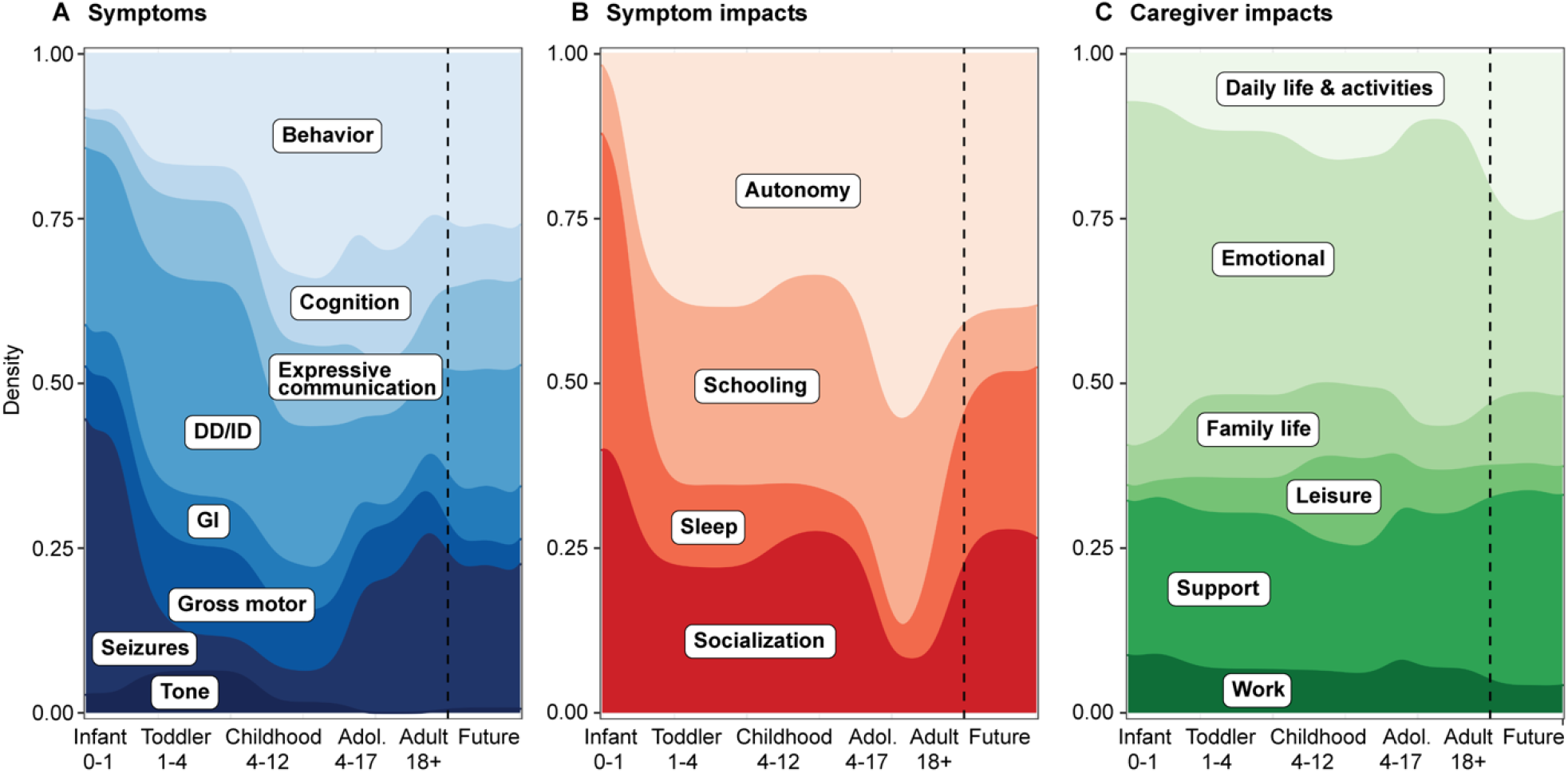
Distribution of symptoms, symptom impacts, and caregiver impacts across the life span in *STXBP1*-related disorders. Concepts were referenced to age groups, including infancy (<12 months), toddlerhood (1-4 years), childhood (5-12 years), teenage & adolescence (13-17 years), adulthood (18+ years), and future considerations. **(A)** Relative density of the most frequently referenced symptoms. **(B)** Relative density of the most frequently referenced symptom impacts. **(C)** Relative density of the most frequently referenced caregiver impacts.

**Figure 5.**
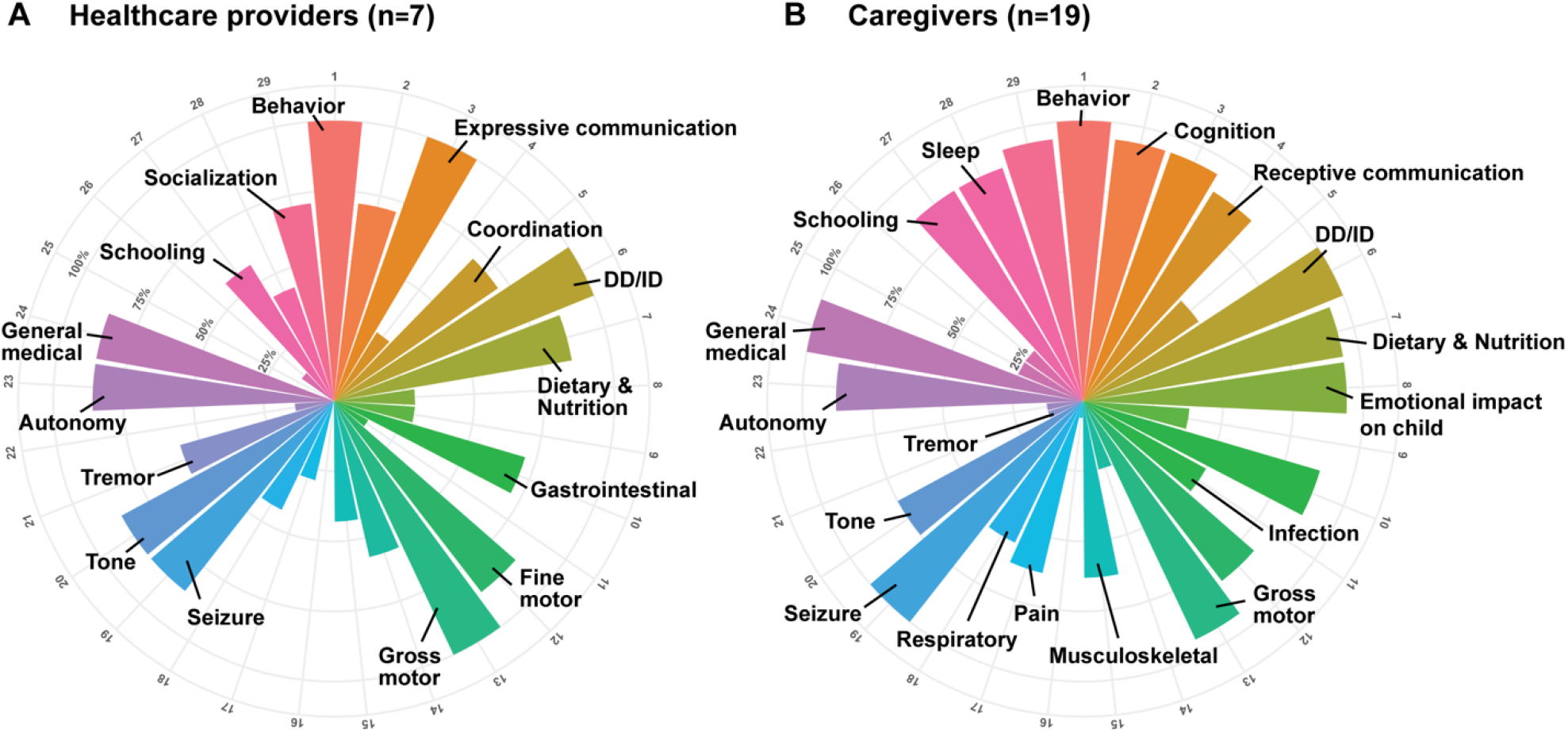
Proportion of interviews in which concepts were referenced at least once by healthcare providers or caregivers. **(A)** Distribution of concepts in the symptoms and symptom impacts domains across healthcare providers. **(B)** Distribution of concepts in the symptoms and symptom impacts domains across caregivers. We found that healthcare providers were significantly less likely to reference behavior and more likely to mention developmental delay and movement disorders compared to caregivers of individuals with *STXBP1*-related disorders.

We also identified symptoms underrepresented in the literature including gastrointestinal symptoms (68 references, across 87.5% of interviews), pain (24 references, across 56.3% of interviews) and respiratory symptoms (30 references, across 62.5% of interviews) **(Figure 1 & 5)**. In our previously published cohort of 534 individuals across 19,973 harmonized clinical terms, respiratory symptoms were reported only in 33 individuals (6.2%), gastrointestinal symptoms in 31 individuals (5.8%), and pain in nine individuals (1.7%) [3].

### Symptom Impact (Domain 2): Caregivers referenced autonomy, socialization, and schooling as key symptom impacts on individuals with *STXBP1*-related disorders

We found symptom impacts play an important role in the daily lives of families with *STXBP1*-related disorders, covering almost 66% of the interview time. Ninety-six references regarding the child’s autonomy (87.5% across all interviews), followed by socialization (n=64, 93.8% across all interviews) and schooling (n=61, 87.5% across all interviews) were recorded **(Figure 1 & 5)**. Impacts to the child’s autonomy were frequently double coded with developmental delay (n=52), caregivers’ emotions (n=52), and medical interaction (n=26, **Figure 2**). Caregivers described autonomy impacts as negative in 61%, positive in 18%, and neutral in 21% of cases **(Supplementary Figure 1)**. In contrast, socialization impacts were categorized as positive in 68% and negative in 32% of cases and frequently double coded with behavior (n=27), developmental delay (n=18), and caregivers’ emotions (n=12). Impacts to schooling were frequently double coded with developmental delay (n=27), medical interaction (n=23), caregiver impacts to emotions (n=20), and impacts to daily life & activities (n=16).

### Caregiver Impact (Domain 3): Emotional, support, and daily life & activities concepts reflect major caregiver impacts

Besides the impacts of symptoms on individuals with *STXBP1*-related disorders, we found the impact of *STXBP1* on caregivers is also a major component covering almost 45% of the interview time. Caregivers described emotional, support, and changes to daily life & activities as the main impacts **(Figure 1)**. Emotional impact, support and daily life & activities were mentioned across almost all interviews. Emotional impacts were referenced 354 times and frequently double coded with other concepts, such as: medical interaction (n=144), support (n=94), seizures (n=55), and developmental delays (n=50) **(Figure 2)**. Support, or the participant’s support system, was referenced 200 times across all interviews and was frequently double coded with caregiver impacts to emotions (n=94), medical interaction (n=53), daily life & activities (n=25), and developmental delays (n=22). The category daily life & activities were referenced 108 times across all interviews and was frequently double coded with medical interaction (n=53), developmental delay (n=35), behavior (n=34), and caregiver impacts to emotions (n=27).

### Lived experience for *STXBP1*-related disorders varied across clinical subgroups

While assessing *STXBP1*-related symptoms and their impacts across subgroups stratified by seizure history, ambulation, and verbal communication, we identified important associations that remained significant after correction for multiple testing **(Figure 3)**. Comparing individuals with a history of seizures (n=13), including those with ongoing seizures (n=5) and those in seizure remission (n=8), we found caregivers of individuals with ongoing seizures were five-fold more likely to reference seizures (OR = 4.97, 95% CI 3.41-7.34). They were less likely to reference tone (OR = 0.14, 95% CI 0.02-0.56) and developmental delay (OR = 0.59, 95% CI 0.42-0.82). In contrast, caregivers of individuals in seizure remission were three-fold more likely to reference receptive communication (OR = 3.06, 95% CI 1.36-7.54) and two-fold more likely to reference schooling (OR = 2.24, 95% CI 1.27-4.03).

Comparing ambulatory (n=6) and non-ambulatory (n=9) individuals, caregivers of individuals who were ambulatory were two-fold more likely to reference behavior (OR = 2.31, 95% CI 1.62-3.34) and four-fold more likely to reference pain (OR = 4.5, 95% CI 1.55-17.88). On the other hand, seizures (OR = 0.57, 95% CI 0.38-0.84), visual impairment (OR = 0% CI 0-0.73), respiratory symptoms (OR = 0.22, 95% CI 0.07 -0.58), and dietary & nutrition (OR = 0.4, 95% CI 0.23-0.69) were mentioned less in this subgroup. Caregivers of individuals with verbal communication (n=3) were more likely to reference behavior (OR = 3.05, 95% CI 2.14 -4.34), schooling (OR = 2.43, 95% CI 1.26-4.54), socialization (OR = 2.47, 95% CI 1.3-4.55) and leisure (OR = 2.57, 95% CI 1.43-4.61), and less likely to reference general medical issues (OR = 0.37, 95% CI 0.24-0.58) than caregivers of individuals who were non-verbal.

### Most referenced *STXBP1*-related concepts evolved across the lifespan

To assess the impact of *STXBP1*-related disorders across the lifespan, we associated referenced concepts to predefined age-groups **(Figure 4)**. Sixteen caregivers made references to infancy (<12 months, 542 references), 15 to toddlerhood (1-4 years, 893 references), 19 to childhood (5-12 years, 798 references), four to teenage years (13-17 years, 133 references), four to adulthood (18+ years, 444 references) and 13 to the future (114 references). We found seizures were eight-fold more likely to be referenced related to infancy than any other age category (OR = 8.84, 95% CI 5.97-13.1). General medical issues (OR = 9.10, 95% CI 4.88-18.52), and caregivers’ emotions (OR = 1.84, 95% CI 1.34-2.52) were also more likely to be mentioned within this age group. In toddlerhood, tone (OR = 4.54, 95% CI 1.94-11.49), developmental delay (OR = 1.64 95% CI 1.21-2.21) and fine motor skills (OR = 2.7, 95% CI 1.29-5.78) were referenced more often than in any other age group. In contrast, seizures were less likely to be mentioned in toddlerhood by caregivers (OR = 0.21, 95% CI 0.12-0.37). In the context of childhood, behavior (OR = 2.15, 95% CI 1.56-2.95), schooling (OR = 3.01, 95% CI 1.72-5.26), socialization (OR = 2.26, 95% CI 1.28-3.96) and leisure (OR = 3.34, 95% CI 1.87-5.99) were more likely to be referenced. With respect to teenage years, musculoskeletal symptoms (OR = 6.00, 95% CI 2.10-15.07) and pain (OR = 6.00, 95% CI 2.10-15.07) were six-fold more likely to be referenced. Related to adulthood, infection (OR = 3.99, 95% CI 1.52-10.23), seizures (OR = 1.81, 95% CI 1.19-2.72), sleep (OR = 3.26, 95% CI 1.56-6.68) and daily life and activities (OR = 2.59, 95% CI 1.40-4.64) were overrepresented references compared to other age groups. When referring to considerations for the future, autonomy was seven-fold more likely to be mentioned (OR = 7.17, 95% CI 2.54-20.53).

### Caregivers and healthcare providers differ with respect to symptom references

To assess the impact of *STXBP1*-related symptoms, we also interviewed seven healthcare providers involved in the care of individuals with *STXBP1*-related disorders, complementing the caregivers’ perspectives. While the average interview concept frequency was higher amongst caregivers (73%) than among healthcare professional (51%), the concepts elicited by healthcare professionals largely reflect concepts provided by caregivers. However, some concepts were more likely to be referenced by either caregivers or healthcare providers. Healthcare providers were less likely to reference behavior (OR = 0.37, 95% CI 0.24-0.54) and dietary & nutrition (OR = 0.34, 95% CI 0.15-0.69) and more likely to mention developmental delay (OR = 1.7, 95% CI 1.33-2.18) and movement disorders (OR = 4.21 95% CI 1.61-11.8), including tremors (OR = 5.69, 95% CI 1.81-21.01) **(Figure 5)**.

## Discussion

Characterization of rare disease has historically been based on collections of physician observations, perspectives, and case reports. However, in 1995, Wilson and Cleary modeled a conceptual framework that comprehensively surveyed the entirety of an affected individual or caregiver’s lived experience with complex or rare disease, thereby centering the patients’ experience within the resulting disease definition [18]. Using this model, several studies have created disease concept models (DCMs) of rare diseases, also referred to as *conceptual models, conceptual disease models*, and *patient-centered models*, to ensure patient and caregiver voices contribute to the description of the disorder [14, 15, 19-26]. In addition, regulating entities have endorsed these patient-centered conceptual frameworks and increasingly encouraged drug development to involve patients and caregivers so that their perspectives are prioritized in disease treatment and understanding [27-30].

Here, we report the first DCM for *STXBP1*-related disorders, a group of genetic epilepsies that is increasingly receiving attention due to the potential availability of precision medicine therapies. Building on previously generated DCMs for other neurodevelopmental and epilepsy disorders [14, 15, 25, 26], our study reinforces the need to incorporate diverse caregiver and clinician perspectives to fully describe a rare disease. We enrolled participants who varied in ages, sex, race, ethnicities, and diagnostic odysseys so that our DCM could be generalizable to many individuals with *STXBP1*-related disorders. Whereas previously published models have heavily relied on physician opinions to generate the final DCM model, we sought out other expert multidisciplinary providers who could comment on the diagnosis, medical care, therapy needs, and education of individuals with *STXBP1*-related disorders.

Standard within qualitative research, semi-structured caregiver and family interviews are assessed for recurrent themes that are formally evaluated using a thematic analysis framework [16]. We find the lived experience of families with *STXBP1*-related disorders range across **three major domains**: symptoms of the condition itself, the impact of these symptoms on the individual with *STXBP1*-related disorders, and their impact on caregivers. We find developmental delay, behavior, and seizures are the predominant symptoms in *STXBP1*-related disorders, but the entire range of reported symptoms spans across a total of 22 distinct concepts, including features such as gastrointestinal and respiratory symptoms that have previously been underrepresented in the previous literature.

Our study has three main findings: first, known clinical features in *STXBP1*-related disorders only represent a subset of the lived experience of families and caregivers. In addition to a wide range of reported clinical characteristics, the full impact of the *STXBP1* mutation on individuals with *STXBP1*-related disorders and their families had not been assessed to date. We find the impact of symptoms and the impact on caregivers play an important role in the daily lives of families of individuals with *STXBP1*-related disorders, as 42% of all concepts are related to these two domains. For future clinical trials, identifying appropriate outcome measures across these domains will be critical. Validated measurement models have been developed to capture impacts of various diseases on the daily life of affected families [31, 32]. Furthermore, ongoing work examines the nuances in developmental and epileptic encephalopathies, such as the association of quality of life (QoL) with the proportion of days that are minimally disrupted by seizures [33].

Our second main finding is the prominent difference with regards to concepts across the age range. Caregivers of individuals with *STXBP1*-related disorders frequently cite seizures and general medical issues when referring to infancy compared to other age-groups. In childhood, social aspects such as behavior and leisure become more prominent. When referring to adulthood, symptoms such as seizures and infections were more frequently mentioned. These findings reflect the dynamic nature of seizure onset, offset, and recurrence, and the impact of social aspects of *STXBP1*-related disorders that parallels other neurodevelopmental and epilepsy DCMs [14, 15, 25, 26, 34] where there is a higher impact of developmental delay and behavioral disorders with a decrease in seizure frequency as the affected individual gets older [35]. This variability across the age-span highlights a challenge for DCMs: when lived experience changes across age groups, establishing general instruments to assess symptom and caregiver impacts may be difficult. Further research is needed to determine how this difficulty can be addressed to assure valid frameworks to capture symptom impact and caregiver impact can be reliably assessed across the life span.

The third main finding emerging from our study is the difference between clinical subgroups. We included this analysis in our study as *STXBP1*-related disorders are traditionally considered to have a very broad range of clinical presentation, a feature that sets *STXBP1*-related disorders apart from other conditions where DCMs have been developed. Families of individuals with seizures made significantly more references to seizures than those without epilepsy, which is not surprising given the disruptive role of seizures in the families’ lives and made fewer references to developmental and behavioral symptoms. In contrast, families of individuals who walked independently or communicated verbally made more references to behavior and fewer references to general medical concerns. These findings suggest that the impact of behavior in individuals with *STXBP1*-related disorders may at least partially be dependent on comorbidities and the overall disease burden on an individual. As our study is one of the first DCMs attempting to compare lived experience across disease subgroups, it is currently unclear whether the inverse relationship of overall disease burden with developmental and behavioral concerns is specific to *STXBP1*-related disorders or a more general feature in neurodevelopmental disorders. There is some literature to suggest that caregivers of children with developmental differences who have both greater caregiver needs and difficult behaviors during caregiving activities experience higher levels of stress [36]. Additionally, within pediatric populations with cerebral palsy (CP), research has shown families report greater levels of self-efficacy when caregiving for children with higher gross motor function [37].

Furthermore, we captured a difference between caregivers and healthcare providers. These groups differed with regards to relative references to symptom and symptom impacts. We found healthcare providers made fewer references to behavior and dietary & nutrition domains to caregivers. Other DCMs have demonstrated differences between caregiver and healthcare provider references, which identifies potential discrepancies between these cohorts for treatment prioritization and impact importance. Additionally, when comparing symptoms referenced in our cohort to the disease spectrum captured in a previously published cohort of 534 individuals, we found gastrointestinal symptoms, respiratory symptoms, and pain are underrepresented in clinical documentation despite being consistently mentioned across our interviews [3]. This highlights a potential gap in clinical care that could be addressed by providers who follow individuals with *STXBP1*-related disorders.

Limitations of our study include the limited number of healthcare providers interviewed per subspecialty, number of adults with *STXBP1*-related disorder included, and power to compare between subgroups. While few healthcare providers were interviewed per clinical specialty, this multidisciplinary panel had over 140 years of professional experience and followed approximately 110 individuals with *STXBP1*-related disorders ranging from infancy to adulthood. Our study greatly benefitted from the diverse perspectives of multidisciplinary experts reflective of a clinical team assembled to manage individuals with *STXBP1*-related disorder. We acknowledge that our study needed to be based on caregiver interviews and not the affected person themselves; additionally, while our study population was diverse, it may not represent the experience of all caregivers and health care providers of individuals with *STXBP1*-related disorders. Nevertheless, due to the small number of adult individuals diagnosed with *STXBP1*-related disorders, our cohort is limited to four adult individuals and may not represent all adult histories. We expect the conceptual model of *STXBP1*-related disorders will change as current patients age, and more patients from a wider spectrum of severity are diagnosed in the coming decades. Furthermore, this study reflects the experience of caregivers and providers of individuals with *STXBP1*-related disorders receiving care from a selection of institutions in the United States. Nevertheless, DCMs of Dravet Syndrome, developed with families living in four different countries, showed that the main disease concepts are similar despite different health care systems [14]. For *STXBP1*-related disorders, we also expect similar disease concepts to be referenced by families living outside of the US.

In summary, we developed a disease concept model for *STXBP1*-related disorders containing 38 distinct concepts to present the lived experiences of families with *STXBP1*-related disorders. We captured symptoms and their impact on individuals and their caregivers across different clinical subgroups and age ranges. We found clinical features previously underrepresented in literature and less frequently referenced by health care providers compared to caregivers. This first comprehensive disease model of *STXBP1*-related disorders is essential as a foundation for developing standard of care protocols to identify endpoints in future clinical trials, focusing on the concepts that are important to affected individuals and their caregivers.

## Data Availability

The data that support the findings of this study are available on request to the corresponding author.

## Acknowledgements

We thank the families and individuals with *STXBP1*-related disorders for making this study possible. This study was funded in part by the *STXBP1* Foundation. Individuals from COMBINEDBrain provided essential input guiding study design and data collection efforts.

## Data Availability Statement

The data that support the findings of this study are available on request from the corresponding author.

## IRB Statement

This study was completed per protocol with local approval by the Children’s Hospital of Philadelphia (CHOP) Institutional Review Board (IRB 21-018734).

## Author Contributions

**Katie R Sullivan:** Conceptualization; Data Curation; Formal Analysis; Investigation; Methodology; Software; Validation; Visualization; Writing – Original Draft Preparation; Writing – Review & Editing

**Sarah M Ruggiero:** Conceptualization; Data Curation; Formal Analysis; Funding Acquisition; Investigation; Methodology; Project Administration; Resources; Software; Supervision; Validation; Visualization; Writing – Original Draft Preparation; Writing – Review & Editing

**Julie Xian:** Conceptualization; Data Curation; Formal Analysis; Investigation; Methodology; Software; Validation; Visualization; Writing – Original Draft Preparation; Writing – Review & Editing

**Kim M Thalwitzer:** Conceptualization; Data Curation; Formal Analysis; Investigation; Methodology; Software; Validation; Visualization; Writing – Original Draft Preparation; Writing – Review & Editing

**Sydni Stewart:** Data Curation; Methodology

**Mahgenn Cosico:** Data Curation; Methodology

**Jackie Steinburg:** Conceptualization; Funding Acquisition; Investigation; Validation; Methodology; Writing – Review & Editing

**James Goss:** Conceptualization; Data Curation; Formal Analysis; Funding Acquisition; Investigation; Validation; Methodology; Writing – Review & Editing

**Anna Pfalzer:** Conceptualization; Methodology; Writing – Review & Editing

**Kyle J Horning:** Conceptualization; Methodology; Writing – Review & Editing

**Nicole Weitzel:** Conceptualization; Methodology; Writing – Review & Editing

**Sydney Corey:** Conceptualization; Methodology; Writing – Review & Editing

**Laura Conway:** Supervision; Project Administration; Writing – Review & Editing

**Charlene Son Rigby:** Conceptualization; Data Curation; Formal Analysis; Funding Acquisition; Investigation; Validation; Methodology; Writing – Review & Editing

**Terry Jo Bichell:** Conceptualization; Data Curation; Formal Analysis; Methodology; Supervision; Investigation; Validation; Writing – Review & Editing

**Ingo Helbig:** Conceptualization; Data Curation; Formal Analysis; Funding Acquisition; Investigation; Methodology; Project Administration; Resources; Software; Supervision; Validation; Visualization; Writing – Original Draft Preparation; Writing – Review & Editing

