## Supplementary Material for "A disease concept model for *STXBP1*-related disorders"

**Supplemental Table 1.** Representative caregiver quotes

**Supplemental Figure 1**. Positive and negative impacts for concepts in *STXBP1*-related disorders: socialization, behavior, and autonomy

**Supplemental Figure 2**. Landscape of age-related concept associations across the life span of *STXBP1*-related disorders

**Supplemental File 1**. Caregiver interview guide for *STXBP1*-related disorders disease concept model

**Supplemental File 2**. Physician & Genetic Counselor interview guide for *STXBP1*-related disorders disease concept model

**Supplemental File 3**. Physical Therapist & Occupational Therapist interview guide for *STXBP1*-related disorders disease concept model

**Supplemental File 4**. Educator interview guide for *STXBP1*-related disorders disease concept model

**Supplemental File 5**. Seizure checklist guide for *STXBP1*-related disorders disease concept model

**Supplemental File 6**. Symptom patient-impact checklist guide for *STXBP1*-related disorders disease concept model

**Supplemental File 7.** Symptom caregiver-impact checklist guide for *STXBP1*-related disorders disease concept model

**Supplemental File 8.** NVivo Codebook

**Supplemental Table 1.** Representative caregiver quotes

| **Concept** | **Caregiver’s quote** |
| --- | --- |
| S - Behavior | *‘He has disruptive tendencies where, he takes his bed apart every day, takes the mattress off, takes the sheets and pillows off. He'll knock over things when he's upset or in pain.’* |
| S - Cognition | *‘Um, she kind of knows [her classmates’] names on her talker.’* |
| S - Communication - Expressive | *‘Uh, he's still nonverbal. He uses a facilitated communication device, an iPad with an alphabet display that reaches words for him.’* |
| S - Communication - Receptive | *‘I don’t think she understands conversations, but she, she will understand instructions, that are common. Like ‘go get it’ or ‘give me’ or anything of that sort.’* |
| S - Coordination | *‘But if she’s having cereal or oatmeal in the morning on a spoon, there is just no way with her ataxia so she needs to be fed.’* |
| S - Developmental Delay | *‘Um, he was delayed in sitting, walking, standing. Yeah, like crawling, standing. Everything was delayed, obviously.’* |
| S - Dietary & Nutrition | ‘*We had to take her to [Hospital] and they put her in their feeding program, and within six months, they got her from baby food to actual people food.’* |
| S - Emotional | *‘She’s like the happiest kid. Like she is probably happier than she would be if she were freaking typical. I don’t know any other [0-5-year-old children] that are just, like, just full of sunshine.’* |
| S - Gait | *‘So, you know, walking was delayed, when she walked, she was very rigid.’* |
| S - Gastrointestinal | *‘Oh, yeah. Constipation. Big time. There's a whole, like, pooping ritual we have in our house…’* |
| S - Infection | *‘So because she is in diapers and if she gets diarrhea, it’s very easy to for her to get an UTI.’* |
| S - Motor - Fine Motor | *‘So we work on getting him to eat better and use a fork and utensils. He doesn’t drink from a cup or from a straw.’* |
| S - Motor - Gross Motor | *‘She only sat up at one year old. Like I said, she started crawling at about two. She walked at three…’* |
| S - Movement Disorder | *‘His involuntary movements. Especially when he’s excited.’* |
| S - Musculoskeletal | *‘But she had scoliosis before she had her broken bones.’* |
| S - Neurological (OTHER) | *‘He has headaches occasionally.’* |
| S - Pain | *‘A difficult day for [Child] is when he's in a lot of pain. He has trouble expressing that pain or alleviating it. Uh, he is looking for any kind of relief, and he will just act out, start destroying whatever's in front of him, knock over a dresser.’* |
| S - Respiratory | *‘When he gets sick leads to respiratory infections, respiratory infections lead to vomiting, vomiting leads to aspiration pneumonia. Aspiration pneumonia means hospitalization.’* |
| S - Seizure | *‘So I would say, 99% of her seizures are in and out of sleep, and they're short 10, 10 to 12 seconds.’* |
| S - Tone | *‘He has little muscle tone, especially in his shoulder trunk in his neck area, and I also feel like his esophagus muscles also have low muscle tone because we do have issues with swallowing sometimes.’* |
| S - Tremors | *‘She has tremors in her hands. So they decided, you know, that we try specifically testing for movement disorders.’* |
| S - Visual | *‘Although she wears glasses all the time, and her vision has improved a lot. Initially she has astigmatism. Initially, her number of glass* [sic] *was very high.’* |
| SI - Autonomy | *‘Uh my [child] is [15-20 years old]. She's stable. She does not walk, does not talk. Does not, um, eat or go to the bathroom on her own. Okay, she is basically...she basically needs all things taken care of.’* |
| SI - General Medical/  Medical Interaction | *‘Um, the clinician basically is, like, was looking at the monitor and left the room and called someone immediately because they noticed hypsarrhythmia on the screen or whatever they’re looking at?’* |
| SI - Puberty | *‘Um, and ovulation, the cramping was so severe, and I guess she didn’t know or didn’t understand it.’* |
| SI - Regression | ***‘****At that point he was developing normally and again he was fairly young, too. Um, so they, you know, I think at that point, he started like, like, to hold his head, good eye contact. You know, the little babbles and stuff like that. Then afterwards* [the seizure episode]*, you know, he lost his head control and stuff like that which I know is fairly common.’* [response regarding temporary regression following seizure episode] |
| SI - Schooling | *‘He's in school for a normal full day. So he then, you know, he gets special instructions PT, OT, and speech, you know, adaptive specials. Adapted PE, adaptive art, music.’* |
| SI - Sleep | *‘And that's transitioning from sleep to awake or awake to sleep. So that's a very complex transition for her brain…I would say, 99% of her seizures are in and out of sleep...’* |
| SI - Socialization | *'Yeah, he really loves reaching out to other people. And it doesn't matter if he's ever seen them before a day in his life or not.’* |
| CI - Daily life & activities | ***‘****So a typical day would, she gets up at 5:30 in the morning. And, you know, we get her dressed, we brush her hair. Like I said, we don't brush her teeth because she does not allow it. Um, so, you know, we get her all ready.’* |
| CI - Emotional | *‘When my child had seizures as an infant, it made me scared.’* |
| CI - Family life | *‘Or she sees her siblings around eating. So I think she wants that… So I think she wants to enjoy being around her siblings…’* |
| CI - Financial | *‘I ended up suing the school district, uh, and spent a lot of money out of my own pocket to get him into this.’* |
| CI - Leisure | *‘Um, all of her equipment, all of her medication. Um, it's a lot packed into, you know, just for her. Everyday living is a lot in general, but when you're trying to pack that up, go on a little vacation or do something like that, it's a lot. It's a lot of extra baggage.’* |
| CI - Physical | *‘But also like my back is gonna give out one day.’* |
| CI - Sleep | *‘A few years ago, I lived off of, like, two or three hours of sleep.’* |
| CI - Support | *‘I mean, like they know [our child has] a disability. They just don't deal with it very well.’* |
| CI - Work | *‘I didn't know anything about IEP, and then I'm listening to everything. And I'm like, you know, I think I could do this. I think I want to do this. And so I, uh, yeah, I took the test to be a teacher, and then I got into a credential program and yeah, and became a special teacher [more than 20 years ago].’* |

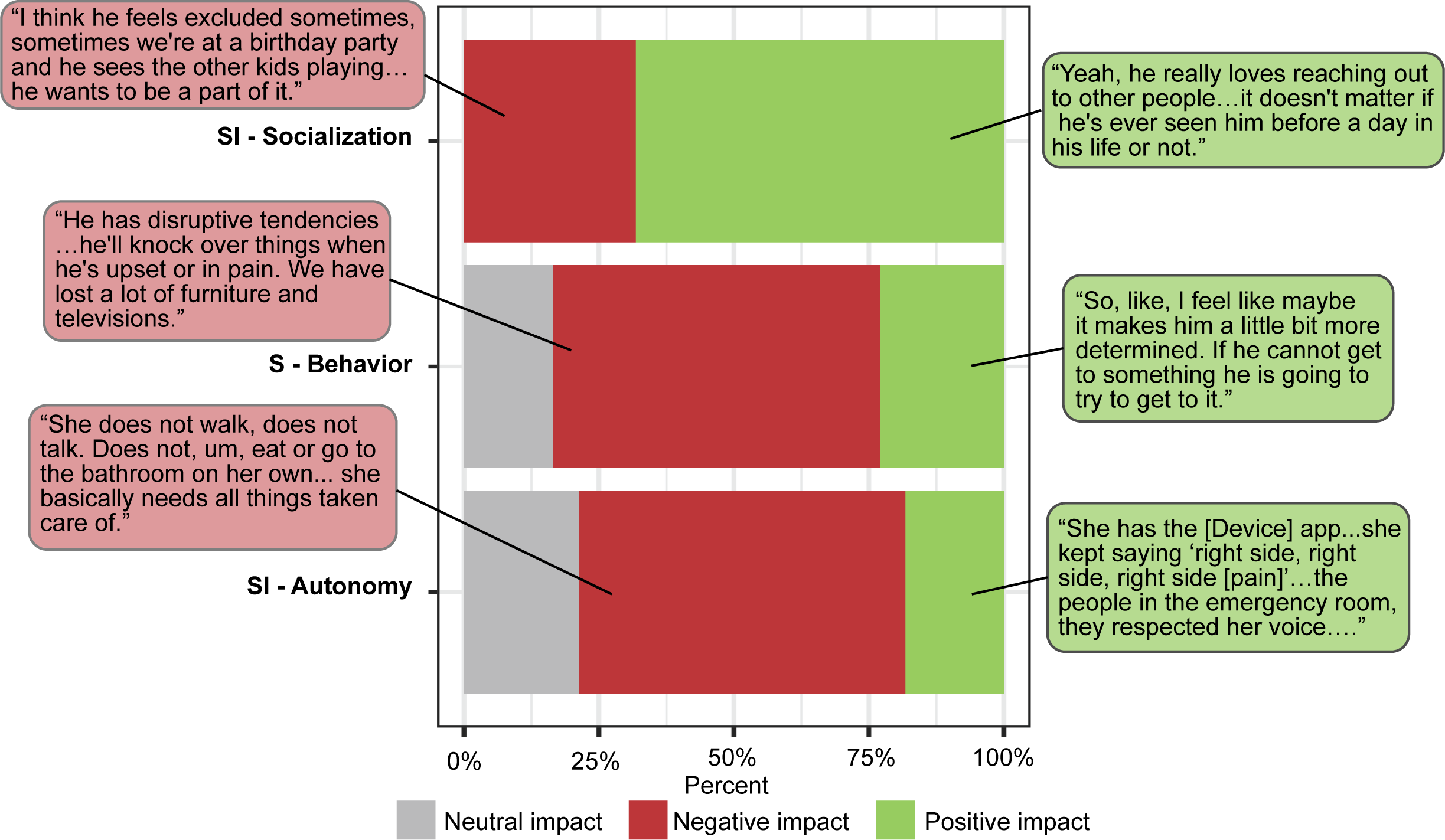

**Supplemental Figure 1**. **Positive and negative impacts of concepts in *STXBP1*-related disorders: socialization, behavior, and autonomy.** For a selection of concepts, we reviewed the direct references in interview transcripts and determined the overall impact (i.e., positive, negative, neutral). Shown here are the relative proportion of positive and negative impacts across the selected concepts, highlighting example interview quotes for each classification.

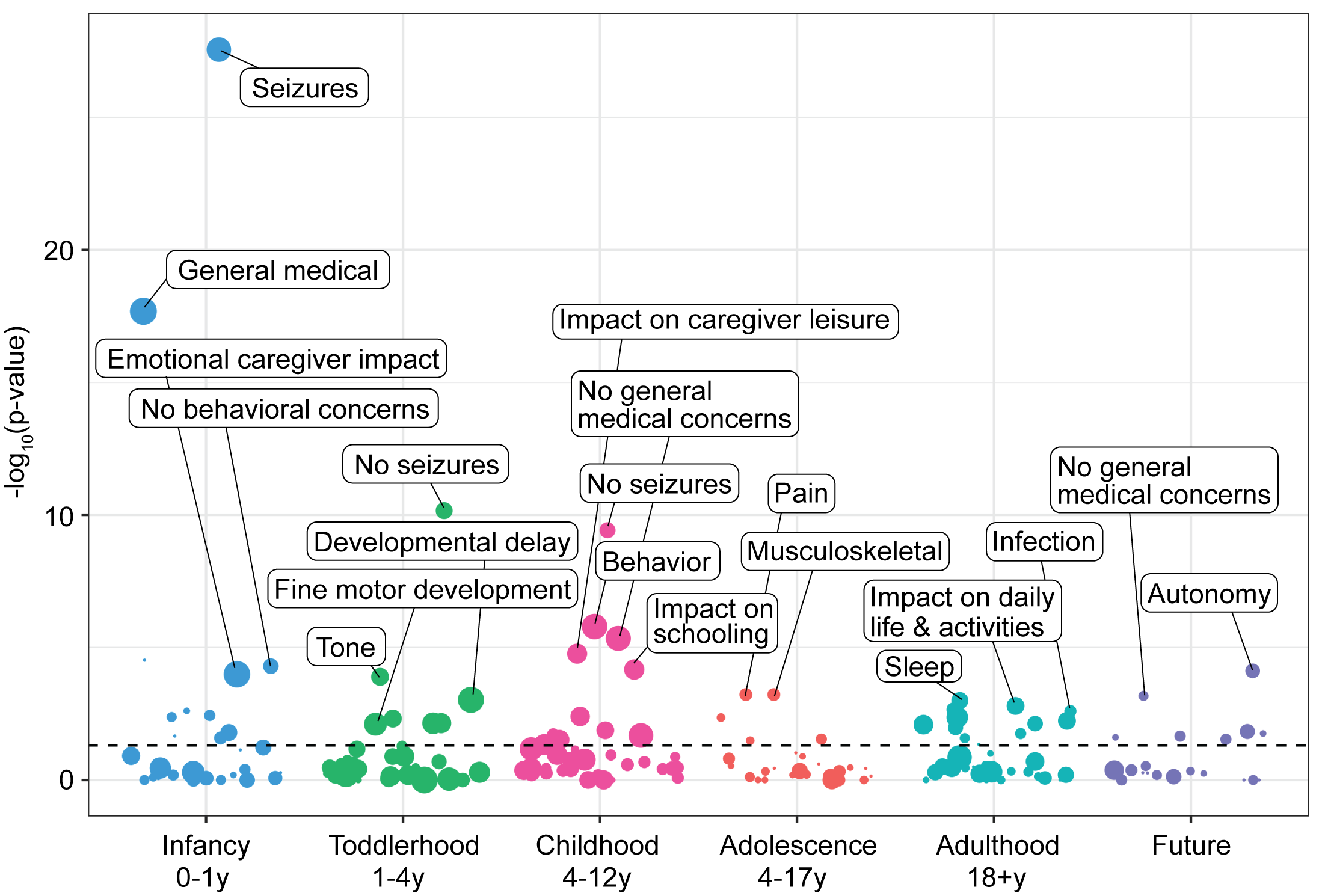

**Supplemental Figure 2**. **Landscape of age-related concept associations across the life span of *STXBP1*-related disorders.** Concepts associated to age groups, infancy (<12 months), toddlerhood (1-4 years), childhood (5-12 years), teenage & adolescence (13-17 years), adulthood (18+ years), and future considerations, with dashed line indicated a nominal significance threshold of p=0.05. Concepts labeled remained significant after correction for multiple testing.

**Supplemental File 1. Caregiver interview guide for *STXBP1*-related disorders disease concept model**

**Interview date:**

**Name of interviewer:**

**Interview Introduction**

- *“My name is [XXX], and I am performing exploratory research regarding patients with STXBP1-RELATED DISORDER to identify the symptoms and impacts associated with this condition. We are collecting information from a group of caregivers and healthcare professionals involved in the care of children with STXBP1-RELATED DISORDER.”*
- *“Today we will talk about how STXBP1-RELATED DISORDER affects your child. We will discuss symptoms your child experiences, how STXBP1-RELATED DISORDER affects their life, the treatments they receive, and the impact of those treatments. This interview will last approximately 60 minutes.”*
- *“Your name and contact information will remain with the Epilepsy Neurogenetics Initiative and will only be accessible to researchers directly involved with this project. We may recontact you if it is necessary to clarify points raised in the interview. Any information you provide will be reported in a way that protects your privacy by avoiding any mention of your name or other information that could identify you.”*
- *“The interview today will be audio-recorded to enable us to pay careful attention to what you say and to make certain we accurately capture the information that you provide to us during the interview. After the interview the audio-recording will be transcribed (written out)”*
- *“Please try to speak relatively loudly so that your comments can be heard and are clear on the recordings.”*
- *“Please be honest in your responses and don’t be afraid to voice any opinions. We want to know about your experience and opinions, not those of your doctor or anyone else.”*
- *“If you do not understand a question, please ask for the question to be repeated and clarified.”*

I**MPORTANT**: *“If you find any of the questions difficult to answer, please let me know. You do not have to answer any questions you do not want to. You may leave the interview at any point you feel the need to, and you may decline to answer any question.”*

**TURN ON THE AUDIO RECORDING and ask the participant the following questions:**

1) ***“Do you agree to participate in this interview?”***
2) ***“Do you agree to have this interview audio-recorded?”***
3) ***“Do you have any questions at this point?”***

**Instructions for the interviewer:**

The following interview guide is designed as a semi-structured format where the interviewer will not ask leading questions but instead explore points that the participant raises through non-leading probes. Some examples of non-leading probes include:

- Can you describe exactly how that feels?
- Tell me more about that.
- How does that affect you?
- Can you talk more about ___________?
- How often does that happen?
- How long does that last?
- Is there anything that makes it better or worse?
- How do you cope with that?
- What makes you say that?

It may not be necessary to ask every question in this guide (i.e., if the participant does not list seizures as a part of their clinical picture, then we do not need to ask questions related to seizure history).

Questions in this guide may be explored in any order.

**Caregiver intake questions:**

1. What is your gender identity?
2. What is your age (years)?
3. What is your race/ethnicity?
4. What is your child’s race/ethnicity?
5. What is your child’s gender?
6. How old is your child (years)?
7. How old was your child when they were diagnosed with STXBP1-related disorder (please specify years or months)?

### **Discussion about STXBP1-related disorder**

### 1. When you talk to your family or friends about what is happening with your child’s health, how do you refer to your child’s condition? What do you call it? Is it ok to use this term throughout the interview?

2. First, I would like to discuss some background information and the symptoms your child experiences because of his/her disorder
   a. When was she/he first diagnosed with STXBP1-RELATED DISORDER and by whom?
      i. AGE:
      ii. Neurologist
      iii. PCP
      iv. Geneticist
      v. Other:
   b. What were the first symptoms that you noticed?
      i. Seizures:
      ii. Delayed milestones (WILL ASK MORE IN NEXT QUESTIONS)
          1. Feeding difficulties
          2. Eye contact
          3. Rolling over
          4. Sitting
          5. Crawling
          6. Walking
          7. Talking
   c. Please tell me about your child’s developmental milestones:
      1. Rolling over
      2. Sitting
      3. Crawling
      4. **CAREFUL:** Walking
      5. **CAREFUL:** Talking
3. What did a typical day look like when your child was a baby?
5. Can you tell me what a typical day is like for your child?
   a. How do you know what your child is feeling? How do you know?
   b. How does your child act? How does your child behave?
6. Can you tell me what a good day looks like for your child?
   a. What is your child able to do on a ‘good’ day?
   b. What might make a ‘good’ day better than any other?
7. Can you tell me what a bad day looks like for your child?
   a. What is your child able to do/not able to do on a ‘bad’ day?
   b. What might make a ‘bad’ day worse than any other?
8. Does your child attend school or a day-care program?
   a. How is your child doing at school or a day-care program?
   b. **CAREFUL:** Does your child frequently have time off school or day care because of illness or medical needs?
   c. **CAREFUL:** How do the school staff manage your child’s seizures?
9. Can you tell me about your child’s relationship with friends/family?
   a. How does your child socialize?
   b. Does your child socialize with children his/her own age?
   c. Can you tell me about how your child participates in school activities?
   d. How does your child socialize with friends outside of school?
10. Tell me how your child’s participation in the community is affected by STXBP1
    a. What activities does your child find difficult to do because of STXBP1-RELATED DISORDER?
    b. **CAREFUL:** Does your child become tired more often than other children due to his/her STXBP1-RELATED DISORDER?
    c. Is your child able to participate in physical activities?
    d. What type of physical activities does your child need assistance with? Tell me about any support devices or aids (if any) your child uses. How do they help them?
11. How does STXBP1-RELATED DISORDER make your child feel? How do you know?
    a. IF CHILD USES WORDS =, Does he/she tell you, or is this something you observe from your child’s behaviors?
12. Have you had to make any adjustments to your home or car to accommodate your child?
    a. What type of adjustments have you made?
    b. Have you had to pay for these adjustments yourself or are reimbursed? If reimbursed, by whom?
13. What changes in your child’s health would make a big difference in his/her daily life?
14. What changes in your child’s health would make a big difference in your daily life?
15. What changes in your child’s health would make a big difference in your family’s daily life?
16. When your child was younger were different symptoms challenging? Have they changed over time? Can you please tell me the symptoms that were the most challenging when your child was:
    a. Less than 5 years of age?
    b. 6-12 years?
    c. 13-18 years?
17. Are there any other examples that you can think of about how STXBP1 has affected your child’s life that we did not ask and that you would like to mention?
18. l would like to understand which symptoms are most challenging to manage. Can you describe the 3 symptoms that your child is experiencing at this time, that are the most challenging to manage as a parent/caregiver
20. Tell me about what **THERAPIES** your child receives
21. What impact does the **THERAPY** have on your child’s life? How do you know?
    a. Have the **THERAPIES** your child receives changed over time? If so, how has it changed?
22. Tell me about which types of **TREATMENT** your child receives

**Parent/caregiver impact of STXBP1-RELATED DISORDER (15 minutes)**

1. What type of care do you do for your child?
2. Can you describe how caring for your child makes you feel?
3. What is the best thing about caring for a child with STXBP1-RELATED DISORDER? Why?
4. What is the hardest thing about caring for a child with STXBP1-RELATED DISORDER? Why?
5. Talk me through a typical day when you are caring for your child. Tell me what it is like from the moment you wake up, to the time you go to bed. What kind of things can you do on a typical day?
a. Is there anything you can’t do on a typical day?
b. How do you feel physically on a typical day?
c. How do you feel emotionally on a typical day?
d. How has this changed since your child was diagnosed with STXBP1-RELATED DISORDER?
e. Does this change depending on what symptoms your child is experiencing?
6. Describe a ‘good’ day when you are caring for your child. What kind of things can you do on a good day?
a. Is there anything you can’t do on a good day?
b. How do you feel physically on a good day?
c. How do you feel emotionally on a good day?
d. What makes a ‘good’ day better than any other day?
e. How has this changed since your child was diagnosed?
7. Describe a ‘bad’ day when you are caring for your child/the child you care for. What kind of things can you do on a bad day?
a. Is there anything you can’t do on a bad day?
b. How do you feel physically on a bad day?
c. How do you feel emotionally on a bad day?
d. What makes a ‘bad’ day worse than any other day?
e. How has this changed since your child was diagnosed?
8. Tell me about any things you find difficult or cannot do because of your child’s STXBP1-RELATED DISORDER?
9. Tell me about any things you avoid doing because of your child’s STXBP1-RELATED DISORDER?
10. What concerns, if any, do you have about your ability to care for your child?
a. You said [x] was a concern, please explain further.
b. How much are you concerned about this?
c. How does this make you feel?
d. What makes it difficult caring for your child? How much is that a problem for you, if at all?
11. What concerns, if any, do you have about the child you care for?
    a. You said [x] was a concern, please explain further.
    b. How much are you concerned about this?
    c. How does this make you feel?
12. **(If seizures are mentioned**) How do they impact daily life? Does this change during the day or night?
13. What, if anything makes caring for your child/the child you care for easier?
    a. How does that make thing easier?
    b. How does that make you feel?
14. Do you feel you need assistance with caring for your child/the child you care for, if any? Why?
15. What type of assistance do you need, and for which type of caring activities, and how often?
16. What, if anything, could make caring for your child/the child you care for more difficult in the future?
    a. How might this make things more difficult?
    b. How might this make you feel?
17. Finally, is there anything else you would like to tell me about in relation to caring for your child/the child you care for that we have not discussed?

**Supplemental File 2. Physician & Genetic Counselor interview guide for *STXBP1*-related disorders disease concept model**

**Interview date:**

**Name of interviewer:**

**Interview Introduction**

- *“My name is [XXX], and I am performing exploratory research regarding patients with STXBP1-RELATED DISORDER to identify the symptoms and impacts associated with this condition. We are collecting information from a group of caregivers and healthcare professionals involved in the care of children with STXBP1-RELATED DISORDER.”*
- *“Today we will talk about how STXBP1-RELATED DISORDER affects your patients. We will discuss symptoms your patients experience, how STXBP1-RELATED DISORDER affects their life, the treatments they receive, and the impact of those treatments. This interview will last approximately 60 minutes.”*
- *“Your name and contact information will remain with the Epilepsy Neurogenetics Initiative and will only be accessible to researchers directly involved with this project. We may recontact you if it is necessary to clarify points raised in the interview. Any information you provide will be reported in a way that protects your privacy by avoiding any mention of your name or other information that could identify you.”*
- *“The interview today will be audio-recorded to enable us to pay careful attention to what you say and to make certain we accurately capture the information that you provide to us during the interview. After the interview the audio-recording will be transcribed (written out)”*
- *“Please try to speak relatively loudly so that your comments can be heard and are clear on the recordings.”*
- *“Please be honest in your responses and don’t be afraid to voice any opinions.”*
- *“If you do not understand a question, please ask for the question to be repeated and clarified.”*

I**MPORTANT**: *“If you find any of the questions difficult to answer, please let me know. You do not have to answer any questions you do not want to. You may leave the interview at any point you feel the need to, and you may decline to answer any question.”*

**TURN ON THE AUDIO RECORDING and ask the participant the following questions:**

1) ***“Do you agree to participate in this interview?”***
2) ***“Do you agree to have this interview audio-recorded?”***
3) ***“Do you have any questions at this point?”***

Instructions for the interviewer:

The following interview guide is designed as a semi-structured format where the interviewer will not ask leading questions but instead explore points that the participant raises through non-leading probes. Some examples of non-leading probes include:

- Can you describe exactly how that feels?
- Tell me more about that.
- How does that affect you?
- Can you talk more about ___________?
- How often does that happen?
- How long does that last?
- Is there anything that makes it better or worse?
- How do you cope with that?
- What makes you say that?

It may not be necessary to ask every question in this guide (i.e., if the participant does not list seizures as a part of their clinical picture, then we do not need to ask questions related to seizure history).

Questions in this guide may be explored in any order.

**Healthcare Professional Intake Questions**

1. What is your gender identity?
2. What is your age (years)?
3. What is your clinical specialty?
4. What is the number of patients with STXBP1-related disorder you see in a typical month?

**Healthcare professional background (PHYSICIANS & GENETIC COUNSELORS)**

1. Please tell me about yourself and the work you currently do.
   a. What is your job title?
   b. Where do you work?
   c. How many years of clinical experience do you have?
2. Which types of patients would you typically see daily?
   a. Do you work mainly with adults or children?
3. Do the patients you see already have a diagnosis of STXBP1-RELATED DISORDER or are they in the process of being diagnosed?

**Background to STXBP1-RELATED DISORDER (PHYSICIANS)**

1. How often do you see or treat patients with STXBP1-RELATED DISORDER?
   a. What age are the patients with STXBP1-RELATED DISORDER that you treat?
   b. How long have you been treating patients with STXBP1-RELATED DISORDER?
2. At what stage of their condition or treatment do you usually see patients with STXBP1-RELATED DISORDER?
3. Please tell me about the symptoms that patients experience because of STXBP1-RELATED DISORDER? How do patients usually present?
   a. What symptoms do you look out for in patients with STXBP1-RELATED DISORDER?
   b. How do these symptoms vary by the age of the patient? If so, how?
4. Are there any symptoms that particularly characterize children with STXBP1-RELATED DISORDER?
5. Please tell me about how symptoms vary between patients of different ages.
a. From your clinical experience, what are the different age ranges where children experience different symptoms related to STXBP1-RELATED DISORDER?
b. Are there any symptoms that characterize younger children with STXBP1-RELATED DISORDER?
c. Are there any symptoms that characterize older children with STXBP1-RELATED DISORDER?
6. What symptoms are most frequently reported by caregivers?
   a. What symptoms, if any, are experienced less frequently?
7. What symptoms are reported as most severe by caregivers?
   a. What symptoms, if any, are reported as being milder?
   b. Does this change depend on the age of the child?
8. Which symptoms cause you greatest concern? How do you evaluate these symptoms?
9. Please tell me about how symptoms may vary between patients.
   a. Tell me about the differing levels of severity between patient experiences.
   b. Do some patients experience more severe symptoms than others?
   c. Please describe the difference between symptoms of different severities.

**Background to STXBP1-RELATED DISORDER (GENETIC COUNSELORS)**

1. How often do you see patients with STXBP1-RELATED DISORDER?
   a. What age are the patients with STXBP1-RELATED DISORDER that you see?
   b. How long have you been seeing patients with STXBP1-RELATED DISORDER?
2. At what stage of their disease or treatment do you usually see patients with STXBP1-RELATED DISORDER?
3. Please tell me about the symptoms that patients experience because of their STXBP1-RELATED DISORDER? How do patients usually present?
4. Are there any symptoms that particularly characterize children with STXBP1-RELATED DISORDER?
5. Please tell me about how symptoms vary between patients of different ages.
6. What symptoms of STXBP1-RELATED DISORDER are most frequently reported by caregivers?
   a. What symptoms, if any, are experienced less frequently?
7. What symptoms of STXBP1-RELATED DISORDER are reported as most severe by caregivers?
   a. What symptoms, if any, are reported as being milder?
   b. Does this change depend on the age of the child?
8. What concerns about STXBP1-RELATED DISORDER are most frequently reported by caregivers?
9. How do caregiver’s concerns evolve over time?

**Questions about seizures caused by STXBP1-RELATED DISORDER (PHYSICIANS)**

1. How do seizures impact patients who have epilepsy?
2. Tell me about the seizure’s children experience due to STXBP1-RELATED DISORDER?
   a. What are the different types of seizures children can experience?
   b. What are the characteristics associated with each seizure type?
   c. Does this vary between children with STXBP1-RELATED DISORDER of different ages?
   d. From your clinical experience, what would you say are the different age ranges where children with STXBP1-RELATED DISORDER may experience different seizures?
3. How often do children with STXBP1-RELATED DISORDER have seizures?
   a. Does this vary between children?
4. When do children usually experience seizures?
   a. Do children usually have seizures while they are awake? Or asleep?
5. How long do the seizures last when they happen?
6. Do children always experience seizures in the same way, or do they vary?
   a. Is there anything that makes seizures more severe?
7. Do the types of seizures experienced by children change over time? If so, how?
   a. Do you know why this is?
8. Does having seizures ever cause children with STXBP1-RELATED DISORDER injuries or other difficulties?
9. Is there anything you think triggers children with STXBP1-RELATED DISORDER seizures?
   a. Bright or patterned lights?
   b. Warm or cold temperatures?
   c. Physical movement/activity?
   d. Noise?
10. Are there emotional or physical changes before a child with STXBP1-RELATED DISORDER has a seizure?
    a. Are children able to predict when they will have a seizure?
11. Do you discuss sudden and unexpected death from epilepsy (SUDEP) with caregivers of individuals with STXBP1-RELATED DISORDER?
    a. If **NO** =, What reasons do you have for not discussing SUDEP with caregivers of individuals with STXBP1-RELATED DISORDER?
    b. If **YES** =, When do you discuss SUDEP with caregivers of individuals with STXBP1-RELATED DISORDER?

**Diagnosis of STXBP1-RELATED DISORDER (PHYSICIANS & GENETIC COUNSELORS)**

1. How do most children with STXBP1-RELATED DISORDER present to care?
2. Can you tell me about the process of diagnosing children with STXBP1-RELATED DISORDER?
   a. At what age are children usually diagnosed with STXBP1-RELATED DISORDER?
   b. What are the initial concerns that lead to you consider a diagnosis of STXBP1-RELATED DISORDER?
3. Tell me about the treatment’s children with STXBP1-RELATED DISORDER usually receive?
   a. How often does the child receive this treatment(s)?
   b. Does treatment vary between children?
   c. How do you decide what treatment to prescribe to each child?
   d. Does treatment change over time? How? What are the reasons for these changes?
4. What do you think are the most important aspects of STXBP1-RELATED DISORDER to treat?
   a. Why is this?
   b. What aspects of STXBP1-RELATED DISORDER do you see as less important to treat?
5. What are your thoughts about the currently available treatments for STXBP1-RELATED DISORDER?
   a. How do you think treatments for STXBP1-RELATED DISORDER can change?
   b. If there was a key symptom or aspect of STXBP1-RELATED DISORDER that could be changed with treatment, what would this be?
6. What different types of doctors/therapists are usually involved in the treatment of children with STXBP1-RELATED DISORDER?
   a. How do the different doctors/therapists work together to provide treatment for each child?
   b. Which doctors/therapists make decisions about changes to the child’s treatment? What role do you play in this process?
   c. Do the different types of doctors/therapists involvement in patient’s treatment change over time?

**Additional symptoms/features associated with STXBP1-RELATED DISORDER (PHYSICIANS & GENETIC COUNSELORS)**

1. Tell me about any difficulties with **walking or movement** children with STXBP1-RELATED DISORDER experience?
   a. What specific difficulties do children have with walking or movement?
   b. At what age does this usually become a concern?
   c. Does this difficulty vary between children?
2. Do children with STXBP1-RELATED DISORDER have any difficulties with their **muscles**?
   a. What specific difficulties do children have communicating?
   b. At what age does this usually become a concern?
   c. Does this difficulty vary between children?
3. Do children with STXBP1-RELATED DISORDER have any difficulties with their **bones**?
   a. What specific difficulties do children have communicating?
   b. At what age does this usually become a concern?
   c. Does this difficulty vary between children?
4. Do children with STXBP1-RELATED DISORDER have any difficulties with their **teeth, mouth, or jaw**?
   a. What specific difficulties do children have communicating?
   b. At what age does this usually become a concern?
   c. Does this difficulty vary between children?
5. Do children with STXBP1-RELATED DISORDER have any difficulties with their **vision or eyes**?
   a. What specific difficulties do children have communicating?
   b. At what age does this usually become a concern?
   c. Does this difficulty vary between children?
6. Tell me about the difficulties children with STXBP1-RELATED DISORDER have with **communication**?
   a. What specific difficulties do children have communicating?
   b. At what age does this usually become a concern?
   c. Does this difficulty vary between children?
7. Tell me about the difficulties children with STXBP1-RELATED DISORDER have with **fine motor skills**?
   a. What specific difficulties do children have with their fine motor abilities?
   b. At what age does this usually become a concern?
   c. Does this difficulty vary between children?
8. Tell me about the difficulties children with STXBP1-RELATED DISORDER have with **attention**?
   a. What specific difficulties do children have with their ability to attend to tasks?
   b. At what age does this usually become a concern?
   c. Does this difficulty vary between children?
9. Tell me about the difficulties children with STXBP1-RELATED DISORDER have with **eating or drinking**?
   a. What specific difficulties do children have with their ability to eat or drink?
   b. At what age does this usually become a concern?
   c. Does this difficulty vary between children?
10. Tell me about the difficulties children with STXBP1-RELATED DISORDER have with **performing personal hygiene or dress themselves?**
    a. What specific difficulties do children have with their ability to performing personal hygiene or dress themselves?
    b. At what age does this usually become a concern?
    c. Does this difficulty vary between children?
11. Tell me about the difficulties children with STXBP1-RELATED DISORDER have with their **sleeping**?
    a. What specific difficulties do children have with their ability to sleep?
    b. At what age does this usually become a concern?
    c. Does this difficulty vary between children?
12. Tell me about the difficulties children with STXBP1-RELATED DISORDER have with their **behaviors**?
    a. What specific difficulties do children have with their behaviors?
    b. At what age does this usually become a concern?
    c. Does this difficulty vary between children?
13. Can you tell me about any **psychological issues** children with STX experience?
    a. What specific psychological problems do children have?
    b. At what age do these problems usually occur?
    c. Do the difficulties vary between children?
14. Are there any other difficulties children with STXBP1-RELATED DISORDER experience that we have not discussed?
15. That is the end of the questions I have for today. Is there anything else associated with STXBP1-RELATED DISORDER or its treatment you would like to discuss?

**Supplemental File 3. Physical Therapist & Occupational Therapist interview guide for *STXBP1*-related disorders disease concept model**

**Interview date:**

**Name of interviewer:**

**Interview Introduction**

- *“My name is [XXX], and I am performing exploratory research regarding patients with STXBP1-RELATED DISORDER to identify the symptoms and impacts associated with this condition. We are collecting information from a group of caregivers and healthcare professionals involved in the care of children with* *STXBP1-RELATED DISORDER.”*
- *“Today we will talk about how STXBP1-RELATED DISORDER affects your patients. We will discuss symptoms your patients experience, how STXBP1-RELATED DISORDER affects their life, the treatments they receive, and the impact of those treatments. This interview will last approximately 60 minutes.”*
- *“Your name and contact information will remain with Epilepsy Neurogenetics Initiative and will only be accessible to researchers directly involved with this project. We may recontact you if it is necessary to clarify points raised in the interview. Any information you provide will be reported in a way that protects your privacy by avoiding any mention of your name or other information that could identify you.”*
- *“The interview today will be audio-recorded to enable us to pay careful attention to what you say and to make certain we accurately capture the information that you provide to us during the interview. After the interview the audio-recording will be transcribed (written out)”*
- *“Please try to speak relatively loudly so that your comments can be heard and are clear on the recordings.”*
- *“Please be honest in your responses and don’t be afraid to voice any opinions.”*
- *“If you do not understand a question, please ask for the question to be repeated and clarified.”*

I**MPORTANT**: *“If you find any of the questions difficult to answer, please let me know. You do not have to answer any questions you do not want to. You may leave the interview at any point you feel the need to, and you may decline to answer any question.”*

**TURN ON THE AUDIO RECORDING and ask the participant the following questions:**

1) ***“Do you agree to participate in this interview?”***
2) ***“Do you agree to have this interview audio-recorded?”***
3) ***“Do you have any questions at this point?”***

**Instructions for the interviewer:**

The following interview guide is designed as a semi-structured format where the interviewer will not ask leading questions but instead explore points that the participant raises through non-leading probes. Some examples of non-leading probes include:

- Can you describe exactly how that feels?
- Tell me more about that.
- How does that affect you?
- Can you talk more about ___________?
- How often does that happen?
- How long does that last?
- Is there anything that makes it better or worse?
- How do you cope with that?
- What makes you say that?

It may not be necessary to ask every question in this guide (i.e., if the participant does not list seizures as a part of their clinical picture, then we do not need to ask questions related to seizure history).

Questions in this guide may be explored in any order.

**Healthcare Professional Intake Questions**

1. What is your gender identity?
2. What is your age (years)?
3. What is your clinical specialty?
4. How long have you been practicing in your clinical specialty?
5. What is the number of patients with STXBP1-related disorder you see in a typical month?

**Healthcare professional background**

1. Please tell me about yourself and the work you currently do.
   a. What is your job title?
   b. Where do you work?
   c. How many years of clinical experience do you have?
2. Which types of patients would you typically see daily?
   a. Do you work mainly with adults or children?
3. Do the patients you see usually already have a diagnosis of STXBP1-RELATED DISORDER or are they in the process of being diagnosed?

**Background to STXBP1-RELATED DISORDER**

1. How often do you see patients with STXBP1-RELATED DISORDER?
   a. What age are the patients with STXBP1-RELATED DISORDER that you see?
   b. How long have you been seeing patients with STXBP1-RELATED DISORDER?
2. At what stage of their disease or treatment do you usually see patients with STXBP1-RELATED DISORDER?
3. Please tell me about the symptoms that patients experience because of their STXBP1-RELATED DISORDER? How do patients usually present?
   a. What symptoms do you look out for in patients with STXBP1-RELATED DISORDER?
   b. How do these symptoms vary by the age of the patient? If so, how?
4. Are there any symptoms that particularly characterize children with STXBP1-RELATED DISORDER?
5. Please tell me about how symptoms vary between patients of different ages.
a. From your clinical experience, what would you say are the different age ranges where children experience different symptoms?
b. Are there any symptoms that characterize younger children with STXBP1-RELATED DISORDER?
c. Are there any symptoms that characterize older children with STXBP1-RELATED DISORDER?
6. What symptoms of STXBP1-RELATED DISORDER are most frequently reported by caregivers?
   a. What symptoms, if any, are experienced less frequently?
7. What symptoms of STXBP1-RELATED DISORDER are reported as most severe by caregivers?
   a. What symptoms, if any, are reported as being milder?
   b. Does this change depend on the age of the child?
8. What concerns of STXBP1-RELATED DISORDER are most frequently reported by caregivers?
9. Which symptoms cause you greatest concern? How do you evaluate these symptoms?
10. Please tell me about how symptoms may vary between patients.
    a. Tell me about the differing levels of severity between patient experiences.
    b. Do some patients experience more severe symptoms than others?
    c. Describe the difference between symptoms of different severities.

**Additional symptoms/features associated with STXBP1-RELATED DISORDER**

#### We are now going to talk about some of the additional symptoms and features children with STXBP1-RELATED DISORDER syndrome may experience.

1. Tell me about any difficulties with **walking or movement** children with STXBP1-RELATED DISORDER experience?
   a. What specific difficulties do children have with walking or movement?
   b. At what age does this usually become a concern?
   c. Does this difficulty vary between children?
2. Do children with STXBP1-RELATED DISORDER have any difficulties with their **muscles**?
   a. What specific difficulties do children have communicating?
   b. At what age does this usually become a concern?
   c. Does this difficulty vary between children?
3. Do children with STXBP1-RELATED DISORDER have any difficulties with their **bones**?
   a. What specific difficulties do children have communicating?
   b. At what age does this usually become a concern?
   c. Does this difficulty vary between children?
4. Do children with STXBP1-RELATED DISORDER have any difficulties with their **teeth, mouth, or jaw**?
   a. What specific difficulties do children have communicating?
   b. At what age does this usually become a concern?
   c. Does this difficulty vary between children?
5. Do children with STXBP1-RELATED DISORDER have any difficulties with their **vision or eyes**?
   a. What specific difficulties do children have communicating?
   b. At what age does this usually become a concern?
   c. Does this difficulty vary between children?
6. Tell me about the difficulties children with STXBP1-RELATED DISORDER have with **communication**?
   a. What specific difficulties do children have communicating?
   b. At what age does this usually become a concern?
   c. Does this difficulty vary between children?
7. Tell me about the difficulties children with STXBP1-RELATED DISORDER have with **fine motor skills**?
   a. What specific difficulties do children have with their fine motor abilities?
   b. At what age does this usually become a concern?
   c. Does this difficulty vary between children?
8. Tell me about the difficulties children with STXBP1-RELATED DISORDER have with **attention**?
   a. What specific difficulties do children have with their ability to attend to tasks?
   b. At what age does this usually become a concern?
   c. Does this difficulty vary between children?
9. Tell me about the difficulties children with STXBP1-RELATED DISORDER have with **eating or drinking**?
   a. What specific difficulties do children have with their ability to eat or drink?
   b. At what age does this usually become a concern?
   c. Does this difficulty vary between children?
10. Tell me about the difficulties children with STXBP1-RELATED DISORDER have with **performing personal hygiene or dress themselves?**
    a. What specific difficulties do children have with their ability to performing personal hygiene or dress themselves?
    b. At what age does this usually become a concern?
    c. Does this difficulty vary between children?
11. Tell me about the difficulties children with STXBP1-RELATED DISORDER have with their **sleeping**?
    a. What specific difficulties do children have with their ability to sleep?
    b. At what age does this usually become a concern?
    c. Does this difficulty vary between children?
12. Tell me about the difficulties children with STXBP1-RELATED DISORDER have with their **behavior**?
    a. What specific difficulties do children have with their behaviour?
    b. At what age does this usually become a concern?
    c. Does this difficulty vary between children?
13. Can you tell me about any **psychological issues** children with STX experience?
    a. What specific psychological problems do children have?
    b. At what age do these problems usually occur?
    c. Do the difficulties vary between children?
14. Are there any other difficulties children with STXBP1-RELATED DISORDER experience that we have not discussed?

**Therapies for children with STXBP1-RELATED DISORDER**

15. Tell me about the therapies children with STXBP1-RELATED DISORDER usually receive?
    a. How often should children with STXBP1-RELATED DISORDER receive this therapy?
    b. Does the therapy vary between children?
    c. How do you decide what therapy to prescribe to each child?
    d. Does therapy change over time? How? What are the reasons for these changes?
16. What do you think are the most important aspects of STXBP1-RELATED DISORDER to treat?
    a. Why is this?
17. What aspects of STXBP1-RELATED DISORDER do you see as less important to treat?
    a. Why is this?
18. What are your thoughts about the currently available therapy for STXBP1-RELATED DISORDER?
    a. How do you think therapies for STXBP1-RELATED DISORDER can change?
    b. If there was a key symptom or aspect of STXBP1-RELATED DISORDER that could be changed with therapy, what would this be?
19. What different types of doctors & therapists are usually involved in the treatment of children with STXBP1-RELATED DISORDER?
    a. How do the different doctors/therapists work together to provide treatment for each child?
    b. Which doctors/therapists make decisions about changes to the child’s treatment? What role do you play in this process?
    c. Do the different types of doctors/therapists involvement in patient’s treatment change over time?
20. That is the end of the questions I have for today. Is there anything else associated with STXBP1-RELATED DISORDER or its treatment you would like to discuss?

**Supplemental File 4. Educator interview guide for *STXBP1*-related disorders disease concept model**

**Interview date:**

**Name of interviewer:**

**Interview Introduction**

- *“My name is [XXX], and I am performing exploratory research regarding students with STXBP1-RELATED DISORDER to identify the symptoms and impacts associated with this condition. We are collecting information from a group of caregivers and healthcare professionals involved in the care of children with STXBP1-RELATED DISORDER.”*
- *“Today we will talk about how STXBP1-RELATED DISORDER affects your students. We will discuss symptoms your students experience, how STXBP1-RELATED DISORDER affects their life, the treatments they receive, and the impact of those treatments. This interview will last approximately 60 minutes.”*
- *“Your name and contact information will remain with the Epilepsy Neurogenetics Initiative and will only be accessible to researchers directly involved with this project. We may recontact you if it is necessary to clarify points raised in the interview. Any information you provide will be reported in a way that protects your privacy by avoiding any mention of your name or other information that could identify you.”*
- *“The interview today will be audio-recorded to enable us to pay careful attention to what you say and to make certain we accurately capture the information that you provide to us during the interview. After the interview the audio-recording will be transcribed (written out)”*
- *“Please try to speak relatively loudly so that your comments can be heard and are clear on the recordings.”*
- *“Please be honest in your responses and don’t be afraid to voice any opinions.”*
- *“If you do not understand a question, please ask for the question to be repeated and clarified.”*

I**MPORTANT**: *“If you find any of the questions difficult to answer, please let me know. You do not have to answer any questions you do not want to. You may leave the interview at any point you feel the need to, and you may decline to answer any question.”*

**TURN ON THE AUDIO RECORDING and ask the participant the following questions:**

1) ***“Do you agree to participate in this interview?”***
2) ***“Do you agree to have this interview audio-recorded?”***
3) ***“Do you have any questions at this point?”***

**Instructions for the interviewer:**

The following interview guide is designed as a semi-structured format where the interviewer will not ask leading questions but instead explore points that the participant raises through non-leading probes. Some examples of non-leading probes include:

- Can you describe exactly how that feels?
- Tell me more about that.
- How does that affect you?
- Can you talk more about ___________?
- How often does that happen?
- How long does that last?
- Is there anything that makes it better or worse?
- How do you cope with that?
- What makes you say that?

It may not be necessary to ask every question in this guide (i.e., if the participant does not list seizures as a part of their educational picture, then we do not need to ask questions related to seizure history).

Questions in this guide may be explored in any order.

**Educator Intake Questions**

1. What is your gender identity?
2. What is your age (years)?
3. What is your education specialty?
4. How many years of educational experience do you have?
5. What is the number of students with STXBP1-related disorders you have educated?

**Educator background**

1. Please tell me about yourself and the work you currently do.
   a. What is your job title?
2. Which types of students would you typically see daily?
   a. Do you work mainly with adults or children?

**Background to STXBP1-RELATED DISORDER**

1. What is the age of the students with STXBP1-RELATED DISORDER that you educate?
2. How long have you been educating students with STXBP1-RELATED DISORDER?
3. Please tell me about the symptoms that students experience because of their STXBP1-RELATED DISORDER?
4. Are there any symptoms that particularly characterize children with STXBP1-RELATED DISORDER? (From other students)
5. Typical day in the classroom for children with STX?
6. Difficult or challenging day in the classroom for children with STX?
7. What are the most and least important considerations for making a lesson plan for students with STXBP1-RELATED DISORDER?
8. What are the most and least important considerations for making an IEP for students with STXBP1-RELATED DISORDER?
9. What educational concerns or general symptoms of STXBP1-RELATED DISORDER are most frequently reported by caregivers or parents?
   a. What concerns, if any, are experienced less frequently?
10. What educational concerns or general symptoms of STXBP1-RELATED DISORDER are reported as most severe by caregivers or parents?
    a. What symptoms, if any, are reported as being milder?
    b. Does this change depend on the age of the child?
11. What educational concerns or general symptoms cause you the greatest concern? How do you evaluate these symptoms?
12. What do you think are the most important educational aspects of STXBP1-RELATED DISORDER to address?
13. Tell me about the therapies children with STXBP1-RELATED DISORDER usually receive?
    a. How often should children with STXBP1-RELATED DISORDER receive this therapy?
    b. Does the therapy vary between children?
    c. How do you decide what therapy to prescribe to each child?
    d. Does therapy change over time? How? What are the reasons for these changes?
14. How do children with STXBP1-RELATED DISORDER socialize with others in the classroom?
15. Have there been any modifications you have needed to make to your classroom for children with STXBP1-RELATED DISORDER?
16. What is the most challenging part for you with educating a child with STXBP1-RELATED DISORDER?
17. What is the best part for you with educating a child with STXBP1-RELATED DISORDER?
18. Changes to what three symptoms of STXBP1-RELATED DISORDER would improve their educational trajectory over time?

**Additional symptoms/features associated with STXBP1-RELATED DISORDER**

We are now going to talk about some of the additional symptoms and features children with STXBP1-RELATED DISORDER syndrome may experience.

1. Tell me about any difficulties with **walking or movement** children with STXBP1-RELATED DISORDER experience?
   a. What specific difficulties do children have with walking or movement?
   b. At what age does this usually become a concern?
   c. Does this difficulty vary between children?
2. Do children with STXBP1-RELATED DISORDER have any difficulties with their **muscles**?
   a. What specific difficulties do children have communicating?
   b. At what age does this usually become a concern?
   c. Does this difficulty vary between children?
3. Do children with STXBP1-RELATED DISORDER have any difficulties with their **bones**?
   a. What specific difficulties do children have communicating?
   b. At what age does this usually become a concern?
   c. Does this difficulty vary between children?
4. Do children with STXBP1-RELATED DISORDER have any difficulties with their **teeth, mouth, or jaw**?
   a. What specific difficulties do children have communicating?
   b. At what age does this usually become a concern?
   c. Does this difficulty vary between children?
5. Do children with STXBP1-RELATED DISORDER have any difficulties with their **vision or eyes**?
   a. What specific difficulties do children have communicating?
   b. At what age does this usually become a concern?
   c. Does this difficulty vary between children?
6. Tell me about the difficulties children with STXBP1-RELATED DISORDER have with **communication**?
   a. What specific difficulties do children have communicating?
   b. At what age does this usually become a concern?
   c. Does this difficulty vary between children?
7. Tell me about the difficulties children with STXBP1-RELATED DISORDER have with **fine motor skills**?
   a. What specific difficulties do children have with their fine motor abilities?
   b. At what age does this usually become a concern?
   c. Does this difficulty vary between children?
8. Tell me about the difficulties children with STXBP1-RELATED DISORDER have with **attention**?
   a. What specific difficulties do children have with their ability to attend to tasks?
   b. At what age does this usually become a concern?
   c. Does this difficulty vary between children?
9. Tell me about the difficulties children with STXBP1-RELATED DISORDER have with **eating or drinking**?
   a. What specific difficulties do children have with their ability to eat or drink?
   b. At what age does this usually become a concern?
   c. Does this difficulty vary between children?
10. Tell me about the difficulties children with STXBP1-RELATED DISORDER have with **performing personal hygiene or dress themselves?**
    a. What specific difficulties do children have with their ability to performing personal hygiene or dress themselves?
    b. At what age does this usually become a concern?
    c. Does this difficulty vary between children?
11. Tell me about the difficulties children with STXBP1-RELATED DISORDER have with their **sleeping**?
    a. What specific difficulties do children have with their ability to sleep?
    b. At what age does this usually become a concern?
    c. Does this difficulty vary between children?
12. Tell me about the difficulties children with STXBP1-RELATED DISORDER have with their **behaviors**?
    a. What specific difficulties do children have with their behaviors?
    b. At what age does this usually become a concern?
    c. Does this difficulty vary between children?
13. Can you tell me about any **psychological issues** children with STX experience?
    a. What specific psychological problems do children have?
    b. At what age do these problems usually occur?
    c. Do the difficulties vary between children?
14. Are there any other difficulties children with STXBP1-RELATED DISORDER experience that we have not discussed?

**Therapies for children with STXBP1-RELATED DISORDER**

1. Tell me about the therapies children with STXBP1-RELATED DISORDER usually receive?
   a. How often should children with STXBP1-RELATED DISORDER receive this therapy?
   b. Does the therapy vary between children?
   c. How do you decide what therapy to prescribe to each child?
   d. Does therapy change over time? How? What are the reasons for these changes?
2. What do you think are the most important aspects of STXBP1-RELATED DISORDER to treat?
   a. Why is this?
3. What aspects of STXBP1-RELATED DISORDER do you see as less important to treat?
   a. Why is this?
4. What are your thoughts about the currently available therapy for STXBP1-RELATED DISORDER?
   a. How do you think therapies for STXBP1-RELATED DISORDER can change?
   b. If there was a key symptom or aspect of STXBP1-RELATED DISORDER that could be changed with therapy, what would this be?
5. What different types of doctors & therapists are usually involved in the treatment of children with STXBP1-RELATED DISORDER?
   a. How do the different doctors/therapists work together to provide treatment for each child?
   b. Which doctors/therapists make decisions about changes to the child’s treatment? What role do you play in this process?
   c. Do the different types of doctors/therapists involvement in patient’s treatment change over time?
6. That is the end of the questions I have for today. Is there anything else associated with STXBP1-RELATED DISORDER or its treatment you would like to discuss?

**Supplemental File 5. Seizure checklist guide for *STXBP1*-related disorders disease concept model**

**Seizure Checklist**

1. Can you tell me about the seizures your child experiences because of STXBP1-RELATED DISORDER?
2. At what age did your child’s seizures start?
3. How often does your child have seizures?
4. How long do the seizures last when they happen?
5. Does your child always experience seizures in the same way, or do they vary?
6. When does your child usually have seizures?
   a. Does your child usually have seizures while they are awake?
   b. Does your child usually have seizures while they are asleep?
   c. How long has your child had seizures like this?
   d. Do the seizures change if your child is ill? Do they happen more or less frequently?
   e. Does change in your child’s medication affect how often they have seizures?
7. Has the type of seizures experienced by your child changed at all? How have they changed?
8. Has the type of seizure experienced by your child changed since they were diagnosed?
9. Has having a seizure ever caused your child any injuries or significant health change?
10. How do you manage unexpected seizures? (e.g., unusual seizure, long-lasting seizure)
11. Is there anything you think triggers your child’s seizures?
    b. Bright or patterned lights?
    c. Warm or cold temperatures?
    d. Physical movement or activity?
    e. Noise?
    f. Geometric patterns?
    g. Changes in emotional state?
    h. Tiredness?
12. Are you or your child able to predict when they will have a seizure?
13. Does your child take medication for their seizures? If so, how many medications are they on?
    a. How many medications have you discontinued?
14. Has your child experienced any periods of seizure-freedom? How long have they lasted?
15. Has any of your child’s seizure-types stopped completely?
16. Is there anything else you would like to tell me about in relation to your child’s seizures that we have not discussed?

**Supplemental File 6. Symptom patient-impact checklist guide for *STXBP1*-related disorders disease concept model**

| **DOMAIN** | **PATIENT IMPACT** |
| --- | --- |
| ☐ Motor | ☐ Can your child move or walk independently?  ☐ How do they move?  ☐ Can they move independently inside your home?  ☐ Can they move independently outside in the community?  ☐ **IF YES =** Do they have any issues or challenges with walking?  ☐ **IF NO** = Can you describe how your child moves or what they use to move around? |
| ☐ Neurological | ☐ Does your child have difficulty moving their arms and legs?  ☐ **IF YES =** Does your child have a movement disorder? |
| ☐ Muscle/Bones/Teeth | ☐ Low tone/hypotonia  ☐ Bruxism/teeth grinding |
| ☐ Visual | ☐ Does your child look at you directly?  ☐ Does he/she seem to look to the side, rather than straight ahead? |
| ☐ Communication | ☐ Can your child understand verbal instructions?  ☐ How does your child communicate what they need (i.e., if they are hungry or thirsty, or need to use the bathroom)?  ☐ Does your child have any words to communicate verbally? (**If YES**, how many words do they use?)  ☐ Does your child use signs or gestures or communicate?  ☐ How many people can your child communicate with?  ☐ How many people can understand your child’s way of communication?  ☐ Does your child use any devices to aid in communication? |
| ☐ Cognition/attention | ☐ Able to focus for a period (on you or other children?) |
| ☐ Behavior | ☐ Exhibit repetitive or obsessive behavior? |
| ☐ Socializing | ☐ Any difficulty socializing with other children his/her age |
| ☐ GI | ☐ How does your child eat or drink?  ☐ Does your child use a G-tube?  ☐ Does your child’s appetite change at all? Does your child’s appetite change due to their treatment?  ☐ Does your child have any difficulties with digestion? Such as indigestion, constipation, or diarrhea? |
| ☐ Sleeping | ☐ Do you have to make any adjustments so your child will sleep?  ☐ Have any difficulties waking up following sleep?  ☐ What does a typical night look like? A difficult night? |
| ☐ Emotion/psych |  |
| ☐ Feeding, dressing, going to the bathroom |  |
| ☐ Seizures | **(See Seizure Checklist)** |

**Supplemental File 7. Symptom caregiver-impact checklist guide for *STXBP1*-related disorders disease concept model**

| **DOMAIN** | **REFINEMENT QUESTIONS** | **CAREGIVER IMPACT** |
| --- | --- | --- |
| ☐ Daily Life | ☐ Do you get any support in caring for your child?  ☐ What aspects of daily living are affected and how? | ☐ How has this changed since your child was diagnosed?  ☐ Does this differ depending on the symptoms your child is experiencing? |
| ☐ Activities | ☐ What activities are affected and how?  ☐ Are your social activities affected?  ☐ Are you able to go on vacation as a family?  ☐ Have your vacation-making decisions been affected in any way? | ☐ How has this changed since your child was diagnosed?  ☐ Does this differ depending on the symptoms your child is experiencing? |
| ☐ Work | ☐ How has your ability to work (paid work) been affected?  ☐ Has this affected you financially? | ☐ How has this changed since your child was diagnosed?  ☐ Does this differ depending on the symptoms your child is experiencing? |
| ☐ Relationships | ☐ What about your relationships with: partner/family/children/friends/work colleagues?  ☐ How does that make you feel? | ☐ How has this changed since your child was diagnosed?  ☐ Does this differ depending on the symptoms your child is experiencing? |
| ☐ Emotionally | ☐ What feelings/emotions do you have about had or caring for a child with STXBP1-RELATED DISORDER?  ☐ How else can you describe that feeling; what other words might you use to talk about that?  ☐ Are there symptoms that cause you worry or concern? | ☐ How has this changed since your child was diagnosed?  ☐ Does this differ depending on the symptoms your child is experiencing? |
| ☐ Physically | ☐ Can you describe how you are affected physically by your child’s condition? (e.g. back pain from lifting child, tiring from lack of sleep, weight gain from less activity)  ☐ What activities do you avoid/stop doing/do less well because of caring for your child?  ☐ How does that make you feel? | ☐ How has this changed since your child was diagnosed?  ☐ Does this differ depending on the symptoms your child is experiencing? |
| ☐ Sleep | ☐ Can you describe how your sleep is affected?  ☐ Can you tell me about the quality of your sleep?  ☐ How often is your sleep affected?  ☐ Where does your child usually sleep? (e.g., sleep separately or together)  ☐ Does your child sleep through the night? | ☐ How has this changed since your child was diagnosed?  ☐ Does this differ depending on the symptoms your child is experiencing?  ☐ Are there ever any difficulties that arise during the nighttime? |

**Supplemental File 8. NVivo Codebook**

| Name | Description |
| --- | --- |
| Age - Adulthood | Participants describe a period for their child after 18+ years |
| Age - Childhood | Participants describe a period for their child between 5-12 years old |
| Age - Future | Participants describe a period older than their child is currently |
| Age - Infant | Participants describe a period for their child under 1 year old |
| Age - Teenage & Adolescence | Participants describe a period for their child between 13-17 years old |
| Age - Toddler | Participants describe a period for their child between 1-4 years old |
| CI - Daily life & activities | Participants indicate an impact their daily life and activities |
| CI - Emotional | Participants indicate an impact to their own emotions |
| CIE - Negative | Participants indicate an impact to their own emotions that has a negative quality |
| CIE - Positive | Participants indicate an impact to their own emotions that has a positive quality |
| CI - Family life | Participants indicate an impact to their family life |
| CI - Financial | Participants indicate a financial impact |
| CI - Leisure | Participants indicate an impact to their leisure activities |
| CI - Physical | Participants indicate an impact to their physical health |
| CI - Sleep | Participants indicate an impact to their sleep |
| CI - Support | Participants indicate impacts of support **SPECIFY TYPE OF SUPPORT |
| CIS - Community | e.g., social media sites, STXBP1 foundations, rare disease foundations, their immediate community/town/city |
| CIS - Family | Family members |
| CIS - Friend | Friends of the family |
| CIS - No support | Participants explicitly mention having no support system |
| CIS - Nursing | Private nursing staff |
| CIS - Other | Nanny, Social Work |
| CIS - School | School district |
| CIS – Sibling or additional child | Sibling or additional child in the household |
| CIS - Spouse | Current or former spouses, partners |
| CIS - Therapy | Therapists |
| CIS - Work | Employees, bosses, companies |
| CI - Work | Participants indicate impacts to their work **NOT RELATED TO SUPPORT, WHICH WOULD BE CI-SUPPORT>WORK SUPPORT |
| S - Behavior | Participants indicate their child experiences impacts to behavior |
| SB - Negative | Participants indicate their child experiences impacts to behavior that have a negative quality |
| SB - Positive | Participants indicate their child experiences impacts to behavior that have a positive quality |
| S - Cognition | Participants indicate their child experiences impacts to cognition |
| S - Communication - Expressive | Participants indicate their child experiences impacts to expressive communication |
| S - Communication - Receptive | Participants indicate their child experiences impacts to receptive communication |
| S - Coordination | Participants indicate their child experiences impacts to their coordination |
| S - Developmental Delay | Participant describes developmental delays, such as: gross motor, fine motor, expressive language, receptive language, play skills, self-care skills **DOUBLE CODE WITH OTHER SYMPTOM IMPACTS |
| S - Dietary & Nutrition | Participants indicate their child experiences impacts to their diet and nutrition |
| S - Emotional | Participants indicate their child experiences impacts to their emotions |
| S - Gait | Participants indicate their child experiences impacts to their gait **MAY NEED TO BE DOUBLE-CODED WITH SI-COORDINATION |
| S - Gastrointestinal | Participants indicate their child experiences impacts to their gastrointestinal system |
| S - Infection | Participants indicate their child experiences impacts to their immune system or has frequenct infections |
| S - Motor - Fine Motor | Participants indicate their child experiences impacts to their fine motor skills |
| S - Motor - Gross Motor | Participants indicate their child experiences impacts to their gross motor skills |
| S - Movement Disorder | Participants indicate their child experiences movement disorders **DOUBLE CODE WITH OTHER SYMPTOM IMPACTS |
| S - Musculoskeletal | Participants indicate their child experiences impacts to their musculoskeletal system |
| S - Neurological (OTHER) | Participants indicate their child experiences impacts to their neurological system **DOES NOT DESCRIBE SEIZURES OR MOVEMENT DISORDER |
| S - Pain | Participants indicate their child experiences pain |
| S - Respiratory | Participants indicate their child experiences impacts to their respiratory system |
| S - Seizure | Participants indicate their child has experienced seizures. **Includes neonatal seizures, infantile spasms, childhood onset seizures, seizure remission, or seizure reoccurrence **DOUBLE CODE WITH OTHER SYMPTOM IMPACTS |
| S - Tone | Participants indicate their child experiences tone differences **DOUBLE CODE WITH OTHER SYMPTOM IMPACTS |
| S - Tremors | Participants indicate their child experiences tremors |
| S - Visual | Participants indicate their child experiences impacts to their vision |
| SI - Autonomy | Participants indicate their child experiences impacts to their autonomy and general physical functioning, such as: independence, being understood by strangers, expressing needs or wants, recognition of danger and safety awareness |
| SIA - Negative | Participants indicate their child experiences impacts to their autonomy and general physical functioning, such as: independence, being understood by strangers, expressing needs or wants, recognition of danger and safety awareness that has a negative quality |
| SIA - Positive | Participants indicate their child experiences impacts to their autonomy and general physical functioning, such as: independence, being understood by strangers, expressing needs or wants, recognition of danger and safety awareness that has a positive quality |
| SI - General Medical | Participants indicate their child experiences general medical or systemic impacts |
| SI - Puberty | Participants indicate their child experiences impacts regarding puberty **DOES NOT DESCRIBE FEAR OF EVENTUAL PUBERTY, CODE TO CI - EMOTIONAL |
| SI - Regression | Participants express that their child has experienced regression ****DOES NOT DESCRIBE FEAR OR CONCERN FOR REGRESSION, CODE TO CI - EMOTIONAL |
| SI - Schooling | Participants indicate their child experiences impacts pertaining to their schooling |
| SI - Sleep | Participants indicate their child experiences impacts pertaining to their sleep |
| SI - Socialization | Participants indicate their child experiences impacts pertaining to socialization |
| SIS - Negative | Participants indicate their child experiences impacts pertaining to socialization that has a negative quality |
| SIS - Positive | Participants indicate their child experiences impacts pertaining to socialization that has a positive quality |
| XXX - Censor | Interview components that need to be censored post-transcription (contains names, dates, identifying information, etc.) |
| XXX - Coping mechanisms | Participants indicate developing coping mechanisms, such as: modifying changes to house or car, developing communication devices |
| XXX - COVID-19 | Participants indicated impacts due to the COVID-19 pandemic |
| XXX - GOOD QUOTES | Quotes for thesis/publication |
| XXX - Perceived Modifying Factors | Anytime a participant says: ‘at least we don’t have to deal with XYZ’; ‘families with XYZ have it better/worse’ |
| XXX - STX Best part of being caregiver | See caregiver interview guide |
| XXX - STX Cure | See caregiver interview guide |
| XXX - STX Difficult being a caregiver | See caregiver interview guide |
| XXX - STX Feel (Child) | See caregiver interview guide |
| XXX - STX Feel (Parent) | See caregiver interview guide |
| XXX - Therapy & Early Intervention (PT, OT, ST) | Participants indicate their child has or has had PT, OT, ST, early intervention, etc. |
| XXX - UNSURE OF CODE | Code here if you are unsure of code or if we need a new code |
